# Identifying Body Awareness-Related Brain Network Changes after Cognitive Multisensory Rehabilitation for Neuropathic Pain Relief in Adults with Spinal Cord Injury: Delayed Treatment arm Phase I Randomized Controlled Trial

**DOI:** 10.1101/2023.02.09.23285713

**Authors:** Ann Van de Winckel, Sydney T. Carpentier, Wei Deng, Sara Bottale, Lin Zhang, Timothy Hendrickson, Clas Linnman, Kelvin O. Lim, Bryon A. Mueller, Angela Philippus, Kimberly R. Monden, Rob Wudlick, Ricardo Battaglino, Leslie R. Morse

**Affiliations:** Division of Physical Therapy, Division of Rehabilitation Science, Department of Rehabilitation Medicine, Medical School, University of Minnesota, USA; Division of Rehabilitation Science, Department of Rehabilitation Medicine, Medical School, University of Minnesota, USA; Centro Studi di Riabilitazione Neurocognitiva - Villa Miari (Study Center for Cognitive Multisensory Rehabilitation), Santorso, Italy; Division of Biostatistics, School of Public Health, University of Minnesota, USA; Masonic Institute for the Developing Brain, University of Minnesota; Minnesota Supercomputing Institute, University of Minnesota; Department of Physical Medicine and Rehabilitation, Spaulding Rehabilitation Hospital, Harvard Medical School, Boston, Massachusetts; Department of Psychiatry and Behavioral Sciences, Medical School, University of Minnesota, USA; Department of Rehabilitation Medicine, Medical School, University of Minnesota, USA

## Abstract

**Background:** Neuropathic pain after spinal cord injury (SCI) is notoriously hard to treat. Mechanisms of neuropathic pain are unclear, which makes finding effective treatments challenging. Prior studies have shown that adults with SCI have body awareness deficits. Recent imaging studies, including ours, point to the parietal operculum and insula as key areas for both pain perception and body awareness. Cognitive multisensory rehabilitation (CMR) is a physical therapy approach that helps improve body awareness for pain reduction and sensorimotor recovery. Based on our prior brain imaging work in CMR in stroke, we hypothesized that improving body awareness through restoring parietal operculum network connectivity leads to neuropathic pain relief and improved sensorimotor and daily life function in adults with SCI. Thus, the objectives of this study were to (1) determine baseline differences in resting-state and task-based functional magnetic resonance imaging (fMRI) brain function in adults with SCI compared to healthy controls and (2) identify changes in brain function and behavioral pain and pain-associated outcomes in adults with SCI after CMR.

**Methods:** Healthy adults underwent a one-time MRI scan and completed questionnaires. We recruited community-dwelling adults with SCI-related neuropathic pain, with complete or incomplete SCI >3 months, and highest neuropathic pain intensity level of >3 on the Numeric Pain Rating Scale (NPRS). Participants with SCI were randomized into two groups, according to a delayed treatment arm phase I randomized controlled trial (RCT): Group A immediately received CMR intervention, 3x/week, 45 min/session, followed by a 6-week and 1-year follow-up. Group B started with a 6-week observation period, then 6 weeks of CMR, and a 1-year follow-up. Highest, average, and lowest neuropathic pain intensity levels were assessed weekly with the NPRS as primary outcome. Other primary outcomes (fMRI resting-state and functional tasks; sensory and motor function with the INSCI AIS exam), as well as secondary outcomes (mood, function, spasms, and other SCI secondary conditions), were assessed at baseline, after the first and second 6-week period. The INSCI AIS exam and questionnaires were repeated at the 1-year follow-up.

**Findings:** Thirty-six healthy adults and 28 adults with SCI were recruited between September 2020 and August 2021, and of those, 31 healthy adults and 26 adults with SCI were enrolled in the study. All 26 participants with SCI completed the intervention and pre-post assessments. There were no study-related adverse events. Participants were 52±15 years of age, and 1-56 years post-SCI. During the observation period, group B did not show any reductions in neuropathic pain and did not have any changes in sensation or motor function (INSCI ASIA exam).

However, both groups experienced a significant reduction in neuropathic pain after the 6-week CMR intervention. Their highest level of *neuropathic pain* of 7.81±1.33 on the NPRS at baseline was reduced to 2.88±2.92 after 6 weeks of CMR. Their change scores were 4.92±2.92 (large effect size Cohen’s *d*=1.68) for highest neuropathic pain, 4.12±2.23 (*d*=1.85) for average neuropathic pain, and 2.31±2.07 (*d*=1.00) for lowest neuropathic pain. Nine participants out of 26 were pain-free after the intervention (34.62%).

The results of the INSCI AIS testing also showed significant improvements in sensation, muscle strength, and function after 6 weeks of CMR. Their INSCI AIS exam increased by 8.81±5.37 points (*d*=1.64) for touch sensation, 7.50±4.89 points (*d*=1.53) for pin prick sensation, and 3.87±2.81 (*d*=1.38) for lower limb muscle strength. Functional improvements after the intervention included improvements in balance for 17 out of 18 participants with balance problems at baseline; improved transfers for all of them and a returned ability to stand upright with minimal assistance in 12 out of 20 participants who were unable to stand at baseline. Those improvements were maintained at the 1-year follow-up.

With regard to brain imaging, we confirmed that the resting-state parietal operculum and insula networks had weaker connections in adults with SCI-related neuropathic pain (n=20) compared to healthy adults (n=28). After CMR, stronger resting-state parietal operculum network connectivity was found in adults with SCI. Also, at baseline, as expected, right toe sensory stimulation elicited less brain activation in adults with SCI (n=22) compared to healthy adults (n=26). However, after CMR, there was increased brain activation in relevant sensorimotor and parietal areas related to pain and mental body representations (i.e., body awareness and visuospatial body maps) during the toe stimulation fMRI task. These brain function improvements aligned with the AIS results of improved touch sensation, including in the feet.

**Interpretation:** Adults with chronic SCI had significant neuropathic pain relief and functional improvements, attributed to the recovery of sensation and movement after CMR. The results indicate the preliminary efficacy of CMR for restoring function in adults with chronic SCI. CMR is easily implementable in current physical therapy practice. These encouraging impressive results pave the way for larger randomized clinical trials aimed at testing the efficacy of CMR to alleviate neuropathic pain in adults with SCI.

**Clinical Trial registration:** ClinicalTrials.gov Identifier: NCT04706208

**Funding:** AIRP2-IND-30: Academic Investment Research Program (AIRP) University of Minnesota School of Medicine. National Center for Advancing Translational Sciences of the National Institutes of Health Award Number UL1TR002494; the Biotechnology Research Center: P41EB015894, the National Institute of Neurological Disorders & Stroke Institutional Center Core Grants to Support Neuroscience Research: P30 NS076408; and theHigh-Performancee Connectome Upgrade for Human 3T MR Scanner: 1S10OD017974.

## 1. INTRODUCTION

There are 18,000 new cases of Spinal Cord Injury (SCI) every year in the United States with a prevalence of about 299,000 Americans with SCI.^1–3^ Increasing evidence shows that adults with SCI with motor and sensory impairments also experience body awareness deficits.^4–15^ Body awareness refers to an attentional focus on and awareness of internal body sensations, including whole-body awareness as well as awareness of body parts in relation to each other, and how they are positioned and move in space.^16,17^ These motor and sensory impairments, and particularly the loss of awareness of where the limbs are in space,^18^ greatly compromise function and mobility.

Body awareness deficits are thought to contribute to the generation or maintenance of chronic neuropathic pain,^4–7,9,19^ which occurs in about 69% of adults with SCI.^20,21^ Even though many pharmacological treatments are available, patients often report limited effects, and some medications, such as opioids, carry an inherent risk of addiction.^20,22–25^ Therefore, new interventions are needed.

Brain research offers insights into neuropathic pain etiology. Because of the spinal cord lesion, the disrupted communication interferes with the correct processing of sensory signals between the body and the brain. This results in altered structural and functional brain activity, notably in the sensorimotor network and in the pain network,^12,26–39^ which may explain why many adults with SCI perceive abnormal pain signals.^26,27,32–35^ Neuroimaging studies have directly addressed SCI-related neuropathic pain.^30,40–43^ However, there is a lack of consensus as to which brain regions and networks are involved in pain in this heterogeneous population.^30,31,36–39^ Nevertheless, there is strong evidence that the parietal operculum (secondary somatosensory cortex, parts OP1/OP4) and the neighboring insula, also called the parieto-insular cortex, are key areas for both body awareness and pain processing.^6,30,32,44–55^

Therefore, we investigated whether Cognitive Multisensory Rehabilitation (CMR)^56^ −a physical therapy treatment, originally developed for stroke rehabilitation− could be used as a treatment for neuropathic pain and possibly sensory and motor recovery after SCI. We did so because our prior work demonstrated that CMR exercises activate OP1/OP4 and insula;^51,53,57^ and that 6 weeks of CMR restored disrupted resting-state parietal operculum connectivity with other brain areas in adults with chronic stroke, in parallel with upper limb sensory and motor recovery lasting at least 1 year.^49^

CMR focuses on recalibrating body awareness via discrimination exercises. For example, patients have to differentiate various positions of the limbs in space or recognize different positions of the limbs relative to other body parts. The effect of CMR on reduced pain and/or improved sensorimotor function has been reported in adults with stroke, and with shoulder impingement syndrome.^49,58–62^ Our recent case study also reported that an adult with cortical blindness and tetraplegia experienced improved visual and sensorimotor function after CMR, more than two years after the ischemic cortical damage.^63^ The possible mechanisms of CMR were previously reported.^64^ In short, according to this model^64^, which is based on our work and that of others,^46,50–53,64–71^ the brain areas in the multimodal integration network, which include OP1/OP4 and insula, are responsible for integrating all the sensory, motor, and visual information to help create a conscious understanding of what is happening in the body (i.e., whole-body awareness). This information is then sent to the posterior parietal cortex, where visuospatial body maps are created and continuously updated to guide movements in real time.^72–77^ Body awareness and visuospatial body maps are jointly called “mental body representations”. CMR is helping restore those mental body representations.

When sensory information is reduced or absent post-SCI, the brain cannot construct these mental body representations correctly. In sum, these mental body representation deficits hinder functional recovery because the brain does not recognize the body’s spatial location to guide movements.^4–6,12–14,18^ Adults who have SCI-related neuropathic pain often have an altered perception related to the size and dimension of the painful and/or sensorimotor-impaired body parts, which justifies the use of CMR to treat neuropathic pain. They also have an altered perception of body weight, pressure, or touch in those body regions. Because neuropathic pain occurs in adults regardless of their level or completeness of SCI, CMR can also be provided to adults with severe sensorimotor impairments after SCI. Therefore, we hypothesized that improving body awareness through restoring parietal operculum network connectivity leads to neuropathic pain relief and improved sensorimotor and daily life function in adults with SCI.

The objectives of this study were to determine baseline differences in resting-state and task-based functional magnetic resonance imaging (fMRI) brain function in adults with SCI compared to healthy controls and to identify changes in brain function and neuropathic pain and sensorimotor function, as well as SCI and pain-associated outcomes in adults with SCI after CMR.

## 2. METHODS

### 2.1 Design

We recruited participants for a delayed treatment arm phase I randomized controlled trial (RCT) from September 1, 2020, until August 31, 2021. We completed the 1-year follow-up (ranging between 11 months and 1 year 7 months) on December 1, 2022. The detailed protocol has been published in Van de Winckel et al. (2022).^78^

The study was conducted in accordance with the principles of the Declaration of Helsinki (2013)^79^. The University of Minnesota (UMN)’s Institutional Review Board approved the study (IRB #STUDY00008476). The CMR intervention and the International Standards for Neurological Classification of SCI American Spinal Cord Injury Association Impairment Scale (INSCI AIS) exam took place at the University of Minnesota in the first author (AVDW)’s Brain Body Mind Lab. MRI scanning was done at the University of Minnesota’s Center for Magnetic Resonance Research. Questionnaires were completed over the secure UMN Zoom conference platform.

We used the M Health/Fairview healthcare recruitment system in the Twin Cities, Minnesota (MN), to send letters to patients with SCI, without direct involvement of the study staff, so that patients could contact the investigators if they were interested in participating in the study. Participants were also recruited in outpatient locations within a newly funded Spinal Cord Injury Model System Center (Minnesota Regional Spinal Cord Injury Model System) and the community. Specifically, we have collaborators within the Minneapolis VA Health Care System including the Minnesota Paralyzed Veterans; M Health Fairview (Minneapolis); MN SCI associations; Regions Hospital; Courage Kenny Rehabilitation Institute; Get Up Stand Up to Cure Paralysis; Unite2Fight Paralysis, and Fit4Recovery, nursing homes, and long-term care facilities.

### 2.2 Participants

We recruited adults who were between 18 and 75 years old, either uninjured or with SCI of >3 months, medically stable with complete or incomplete SCI, and with the highest level of SCI-related neuropathic pain in the prior week >3 out of 10 on the Numeric Pain Rating Scale (NPRS).^80^

We excluded adults with MRI contra-indications; uncontrolled seizures; severe cognitive impairment; severe vision or hearing impairment; ventilator dependency; major medical complications; pregnant women; children; or those who were involved in other rehabilitation programs that would influence outcomes. Although adults with complete tetraplegia after SCI can potentially benefit from CMR, we only included adults who could self-transfer with assistance to facilitate the transfers from the MRI-safe wheelchair to the MRI bed and to ensure the ability to press the call button with the hand or wrist in case of an emergency in the MRI scanner.

### 2.3 Screening

After consenting, the study staff collected demographic information and general health and medical history. We screened participants for cognitive ability with the short version of the Mini-Mental State Examination (MMSE).^81^ The ability to perform kinesthetic imagery was assessed with the Kinesthetic and Visual Imagery questionnaire (KVIQ),^82^ because this type of imagery was used in the MRI tasks and the CMR exercises also rely partly on kinesthetic imagery.

Participants completed the Center for Magnetic Resonance Research prescreening questionnaire and the Operations and Safety Director reviewed redacted medical information and signed off on MRIs for any participant who had surgeries that involved any type of implant (rods, screws, etc).

### 2.4 Procedures

The procedures for healthy adults were a one-time MRI scan (high resolution structural + resting-state + fMRI tasks) and a remote visit over Zoom to complete non-SCI specific questionnaires, such as mood (Patient Health Questionnaire-9, PHQ-9), anxiety, self-efficacy, and body awareness.

The baseline testing for adults with SCI comprised an MRI (high resolution structural + resting-state + fMRI tasks), a Zoom visit with questionnaires, which included questions on neuropathic pain, function, mood, and SCI-related symptoms, and the INSCI AIS exam for sensation and motor function.

As primary outcome, we assessed weekly the highest, average, and lowest neuropathic pain intensity levels with the NPRS. The fMRI resting-state and functional tasks; the INSCI AIS exam, and the questionnaires were all assessed at baseline and after the first and second 6-week periods. The 1-year follow-up included the INSCI AIS exam and questionnaires over Zoom.

All participants continued their routine health care for SCI management and continued taking their neuropathic pain medication as prescribed and/or on an as-needed basis during the study. After the baseline testing, using computer-generated randomization, adults with SCI were allocated into one of two groups: Group A immediately received 6 weeks of CMR, one-on-one, in-person, 3x/week for 45 minutes/session, followed by 6 weeks of standard of care (no therapy) at home. Group B started with a 6-week observation period with standard of care (no therapy) at home. After this observation period, Group B participants received 6 weeks of CMR because, based on our prior recruitment experience, we anticipated that participants recruited and not offered an intervention would likely drop out. The brain imaging analyst and biostatistician were blinded to group allocation. CMR sessions were recorded on video for quality assurance of the content and delivery of the intervention. Adherence to the in-person CMR sessions was monitored through a log sheet. The intervention was estimated at minimal risk because of the low-intensity movements or kinesthetic imagery used in CMR.

### 2.5 Intervention: Cognitive Multisensory Rehabilitation (CMR)

An experienced CMR therapist provided the CMR intervention to all participants. The goal of CMR is to restore awareness of the paralyzed limbs and trunk in space, in order to improve sensorimotor function.^49,83–85^ A maximum of two different exercises per session were given and usually done sitting or lying on the treatment table. If a person could stand, then exercises were sometimes done in standing position as well, e.g., to train the awareness of body weight shifts when transferring body weight from one foot to the other.

Based on the participant’s description and location of the pain, and how the participant perceived the dimensions of his/her/their body, the CMR therapist chose one of the CMR exercises that involved those body parts. Through the exercise, the participant learned to perceive those body parts better and to relate those body parts with the rest of the body. The therapist also verified whether pain perception changed by having the participant be attentive to those body parts during the exercise.

Types of exercises include identifying the leg position in relation to the pelvis and the upper body; identifying the dimension of the legs and pelvis; the sensation of the pelvis as a central body reference; the relationship between the left and right side of the body, or between the pelvis and the feet. An example of an exercise is as follows: the participant has their eyes closed and is sitting on the treatment table. The therapist places 5 small cardboards under the left foot and 3 under the right foot. The success of CMR lies in how the therapist provides strategies to the participant to solve the exercise in order to restore their whole-body awareness and body movements in space. In this example, the therapist may invite the participant to remember a time in the past when the participant was moving their legs during a well-specified action, such as when riding a bike. Once the participant is able to remember such a feeling, the therapist then asks the participant to use this tool to help him/her/them identify which leg is higher. Recalling a well-specified action from the past (that has a specific context, emotions, and a memory of a body movement) can help the participant access a more complete mental body representation because the memory dates from before the SCI when the mental body representations were intact. This tool helps the brain access these stored mental body representations and integrate them into the current situation. More information about CMR exercises has previously been published.^78^

### 2.6 Outcome measures

#### 2.6.1 Clinical assessments (primary and secondary outcomes)

The primary clinical outcome was the highest, average, and lowest neuropathic pain intensity levels, assessed weekly with the NPRS.^80^ Average neuropathic pain was defined as neuropathic pain intensity felt most of the time in the prior week. The other primary clinical outcome was sensory and motor testing with the INSCI AIS exam.^89^

Secondary outcomes included the NINDS-CDE International SCI Pain Basic Data Set Version 2.0.^90^ This data set assesses average nociceptive and neuropathic pain in the prior week, and identification of body locations where the different types of pain were experienced. It also assesses interference of neuropathic pain with mood, activity, and sleep. We assessed spasm frequency and intensity (Penn Spasm Frequency Scale^91^), SCI-secondary conditions,^92,93^ body awareness (Revised Body Awareness Rating Questionnaire),^94^ anxiety (State and Trait Anxiety Inventory),^95^, mood (symptoms of depression with the Patient Health Questionnaire-9, PHQ-9)^96^, and functional performance with the Spinal Cord Injury Functional Index^97^ and the Patient Specific Functional Scale (PSFS)^98^. The Spinal Cord Injury Functional Index^97^ measures functional performance across five domains: basic mobility, self-care, fine motor function, and ambulation,^99^ with separate questions for adults with paraplegia and tetraplegia. For the PSFS, the participants self-identified goals related to activities in daily life that were important to them but that they currently had difficulty with because of the neuropathic pain. The participants rated them at each time point between 0 (unable to do the activity without pain) and 10 (able to do the activity without pain).^98^ The INSCI AIS exam and a limited version of the questionnaires (about intensity levels of neuropathic pain, interference of pain on activity, mood, and sleep, function, and spasms) and were done at the 1-year follow-up.

During the weekly phone call check-ins, participants were asked to provide information about spasm, neuropathic pain medication intake and/or healthcare use, as well as about what they learned about their body during the sessions, and perceived intervention effects. Data was entered into the University of Minnesota’s REDCap database.

#### 2.6.2 MRI assessments (primary outcome)

Participants were scanned for 2 hours on a Siemens 3-T Prisma scanner at the Center for Magnetic Resonance Research (CMRR). Structural MRI acquisition included a T1-weighted magnetization-prepared rapid acquisition with gradient echo (MPRAGE) [repetition time (TR)=2.5s; echo time (TE)=4.5ms; 0.8mm isotropic voxels] and aT_2_ weighted sampling perfection with application-optimized contrasts using different flip angle evolution (SPACE) [TR=3.2s; TE=565ms; 0.8mm isotropic voxels]. The resting-state and task fMRI scans were obtained with a T2*-weighted multiband echo planar acquisition tipped 30 degrees relative to the anterior commissure–posterior commissure (AC-PC) plane as determined by the auto-align software. This acquisition protocol was designed to measure whole-head blood-oxygen-level-dependent (BOLD)–contrast with optimal temporal and spatial resolution and to reduce signal dropout [TR = 0.8 s; TE = 37 ms; flip angle = 55 degrees; 72 slices; multiband factor 8; 2 mm isotropic voxels].

For the resting-state fMRI imagery (12min 10sec), we selected the parietal operculum (parts OP1/OP4) and insula as region-of-interest (ROI) based on their importance in sensorimotor function, pain, and body awareness.^6,30,32,44–55^ Participants maintained eye fixation with a restful mind and were asked to not fall asleep.^86^

There were five fMRI tasks, with auditory instructions provided through headphones. The fMRI tasks have been chosen to generate activation and connectivity maps of the OP1/OP4 and insula networks.

For Task 1 (pain imagery, 8min 15sec), participants were asked to focus with gentle, nonjudgmental awareness on their most painful body region, versus rest.

For Task 2 (kinesthetic imagery of moving the limb, 7min 58sec), participants were asked to imagine the feeling in the hip and leg as if they were moving their most painful leg back and forth while lying down.

For Task 3 (kinesthetic imagery of moving the whole body, 19min 51sec), participants were asked to imagine the feeling of being upright and performing a whole-body movement. After viewing a video demonstration of a whole-body Qigong movement, in which the Qigong master was standing, and rhythmically, gently opening and closing the arms while bending and stretching the knees in concert with breathing in and out, participants focused on the feeling in the body while they imagined doing this movement.

Task 4 was a sensory stimulation task (18min 32sec), in which an investigator was gently brushing the pads of the big toes and thumbs with a small towel. Participants were not made aware of where or when the gentle brushing with the towel would occur. They were asked afterward if and where they sensed any stimulation on the hands or feet. This task tests residual sensory information reaching the somatosensory cortex, as has been shown using similar tasks in 50% of adults with sensory complete SCI.^87,88^ Strengthening of such responses post-therapy would be evidence of gains in signal transmission past the lesion.

For task 5 (shape discrimination with our MRI-compatible robot, 10min 13sec), the robot moves the finger passively along pre-specified shape trajectories while music clips are playing. Depending on the auditory cue, participants focus on one of the tasks and press a button with the other hand to indicate whether the shapes (or music clips) are the same or different. This protocol has been described in previous publications.^50,51,53^ The goal of the robot task was to identify the cognitive processes related to shape discrimination, or in this case, the awareness and discrimination of movements made by the metacarpophalangeal joint of the index finger. This task was chosen because the majority of our participants were able to feel the finger movements and we knew from our earlier studies that it would activate our areas of interest (i.e., OP1/OP4, insula, sensorimotor cortex, areas in the posterior parietal cortex).

### 2.7 Sample size calculation

In a previous pilot study, we found a significant difference in OP1 connectivity to the periaqueductal gray (*p*FWE=0.0126, *T*=3.3) with an effect size of Cohen’s *d*=1.17 when contrasting 29 adults with SCI to 11 uninjured controls. Based on a two-sample t-test, we estimated that with a sample size of 25 per group (i.e., n=25 healthy adults, n=25 adults with SCI), we would have over 98% power to detect the between-group baseline differences between healthy adults and adults with SCI with an effect size of *d*=1.17 at α-level of 0.05.

For the delayed treatment arm phase I RCT, with three within-subject repeated measurements, assuming a moderate level of correlation of 0.5 between repeated measures, the sample size of 25 participants with SCI would have over 90% power to detect a moderate correlation of *r*=0.46 between brain imaging variables and neuropathic pain at a significance level of 0.05.

Of note, because this was the first CMR study in adults with SCI, the effect of CMR on neuropathic pain perception in the brain or on SCI brain network function was not known. In our prior study of eight adults with stroke, we found reduced resting-state parietal operculum network connectivity at baseline and restored connectivity concurrent with increased sensorimotor function after CMR.^49^ Still, direct power calculations were not possible as the present study was focused on neuropathic pain relief in SCI. The present data are thus crucial to determine the effect size to design a larger clinical trial.

### 2.8 Statistical analysis plan

#### 2.8.1 Clinical assessments

All quantitative variables were summarized using descriptive statistics at each time point. All analyses were conducted using standard statistical software including JMP 15.0 and R. Primary analyses were based on the intent-to-treat (ITT) population. Within-group changes (for the observation period) as well as pre-post CMR changes in both groups combined for neuropathic pain, sensorimotor and other clinical measures were tested using paired t-tests. Between-group differences in the first 6-week period (experimental vs control condition) were analyzed with 2-sample t-tests.

#### 2.8.2 MRI assessments

All neuroimaging data were preprocessed through the Human Connectome Project preprocessing pipeline. FMRI data were processed using the conn functional connectivity toolbox with established standardized controls for multiple comparisons (SPM family-wise error correction methods).^20^ Data underwent realignment, scrubbing, artifact detection, and CompCor denoising. Primary analyses focused on OP1/OP4 and insula as ROI for resting-state connectivity and exploratory multivariate pattern analyses. Task-based fMRI underwent the same preprocessing pipeline and was modeled with a general linear model.

Within-group pre-post changes in task-based brain activation and resting-state connectivity (with Fisher’s Z transformation) were tested with paired t-tests and between-group differences of pre-post changes with 2 sample *t*-tests. We tested the effect of CMR on brain imaging outcomes with mixed effects models and correlated errors after adjusting for potential confounders, which included the subject-level effect, time, group indicator, and time-by-group interaction as predictors, and other covariates as appropriate. We used Benjamini–Hochberg’s False Discovery Rate (FDR) correction to control for the overall Type 1 errors for the voxel-level analyses.

## 3. RESULTS

Thirty-six healthy adults and twenty-eight adults with SCI were recruited between September 2020 and August 2021. ***Figure 1*** shows the CONSORT study flow chart and reasons for exclusion. ***Table 1*** shows the demographic and clinical characteristics of adults with SCI-related neuropathic pain (n=26) and healthy adults (n=31) who enrolled in the study.

**Table 1.** Demographic and clinical characteristics of adults with spinal cord injury-related neuropathic pain and healthy adults.

|  | Adults with SCI and Neuropathic Pain (n=26) | Healthy adults (n=31) |
| --- | --- | --- |
| Age, years, mean±SD, range (years) | 51.69±14.50<br>(25-74) | 40.35±17.33<br>(21-69) |
| Sex, n (%) |  |  |
| Male | 19 (73.08) | 9 (29.03) |
| Female | 7 (26.92) | 22 (70.97) |
| Gender Identity, n (%) |  |  |
| Male | 19 (73.08) | 9 (29.03) |
| Female | 7 (26.92) | 22 (70.97) |
| Other | 0 (0.00) | 0 (0.00) |
| Ethnicity, n (%) |  |  |
| Hispanic | 2 (7.69) | 1 (3.23) |
| Non-Hispanic | 24 (92.31) | 30 (96.77) |
| Race, n (%) |  |  |
| White | 24 (92.31) | 27 (87.10) |
| Asian | 1 (3.85) | 2 (6.45) |
| African American/Black | 1 (3.85) | 1 (3.23) |
| Multi-racial | 0 (0.00) | 1 (3.23) |
| Hawaiian or Other Pacific Islander | 0 (0.00) | 0 (0.00) |
| Other | 0 (0.00) | 0 (0.00) |
| Veteran, n (%) |  |  |
| Yes | 0 (0.00) | N/A |
| No | 26 (100.00) |  |
| Location of residence, n (%) |  |  |
| Rural | 3 (11.54) | N/A |
| City or suburb | 23 (88.46) |  |
| Baseline neuropathic pain intensity level, mean±SD |  |  |
| High | 7.81±1.33 | 0.00±0.00 |
| Average | 5.46±1.53 | 0.00±0.00 |
| Low | 2.62±2.16 | 0.00±0.00 |
| Baseline musculoskeletal pain intensity level, mean±SD |  |  |
| Average | 5.32±1.49 | 0.34±0.60 |
| Time the since spinal cord injury, years, mean±SD, range (years) | 17.83±15.65<br>(1-56) | N/A |
| Etiology of SCI, n (%) |  |  |
| Traumatic | 7 (26.92) | N/A |
| Non-traumatic | 19 (73.08) | N/A |
| Spinal cord injury lesion level, n (%) |  |  |
| Cervical | 4 (15.38) | N/A |
| Thoracic | 18 (69.23) | N/A |
| Lumbar | 4 (15.38) | N/A |
| Sacral | 0 (0.00) | N/A |
| AIS level, n (%) |  |  |
| A | 14 (53.84) | N/A |
| B | 3 (11.54) | N/A |
| C | 4 (15.38) | N/A |
| D | 5 (19.23) | N/A |
| Level of impairment |  |  |
| Paraplegia (complete; incomplete), n (%); n (%) | 14 (53.85); 8 (30.77) | N/A |
| Tetraplegia (complete; incomplete), n (%); n (%) | 0 (0.00); 2 (7.69) | N/A |
| Brown Sequard, n (%) | 2 (7.69) | N/A |
| Ambulation, n (%) |  |  |
| Walking without assistance | 2 (7.69) | 31 (100.00) |
| Combined use of walking with and without assistance | 1 (3.85) |  |
| Walking with assistance | 3 (11.54) |  |
| Manual Wheelchair | 12 (46.15) |  |
| Combined use of Manual and Motorized Wheelchair | 3 (11.54) |  |
| Motorized Wheelchair | 5 (19.23) |  |
| Ambulation, mean percent time (SD) |  |  |
| Walking | 8.85 (±1.06) | N/A |
| Walking with assistance | 14.27 (±1.32) |  |
| Manual Wheelchair | 54.38 (±1.86) |  |
| Motorized Wheelchair | 22.50 (±1.53) |  |
| Neuropathic pain medication at baseline, n (%) |  |  |
| Acetaminophen | 2 (7.69) | 0 (0.00) |
| Baclofen | 2 (7.69) | 0 (0.00) |
| CBD | 1 (3.85) | 0 (0.00) |
| Diazepam | 1 (3.85) | 0 (0.00) |
| Duloxetine | 2 (7.69) | 0 (0.00) |
| Gabapentin | 8 (30.77) | 0 (0.00) |
| Hydrocodone | 2 (7.69) | 0 (0.00) |
| Ibuprofen | 4 (15.38) | 0 (0.00) |
| Lidocaine | 1 (3.85) | 0 (0.00) |
| Lyrica | 3 (11.54) | 0 (0.00) |
| Medical Cannabis THC | 1 (3.85) | 0 (0.00) |
| Methadone | 1 (3.85) | 0 (0.00) |
| Modafinil | 1 (3.85) | 0 (0.00) |
| Naproxen | 1 (3.85) | 0 (0.00) |
| Oxybutynin | 1 (3.85) | 0 (0.00) |
| Oxycodone | 3 (11.54) | 0 (0.00) |
| Oxycontin | 1 (3.85) | 0 (0.00) |
| Pregabalin | 3 (11.54) | 0 (0.00) |
| Tizanidine | 2 (7.69) | 0 (0.00) |
| Tramadol | 2 (7.69) | 0 (0.00) |
| Valium | 1 (3.85) | 0 (0.00) |

**Fig. 1.**
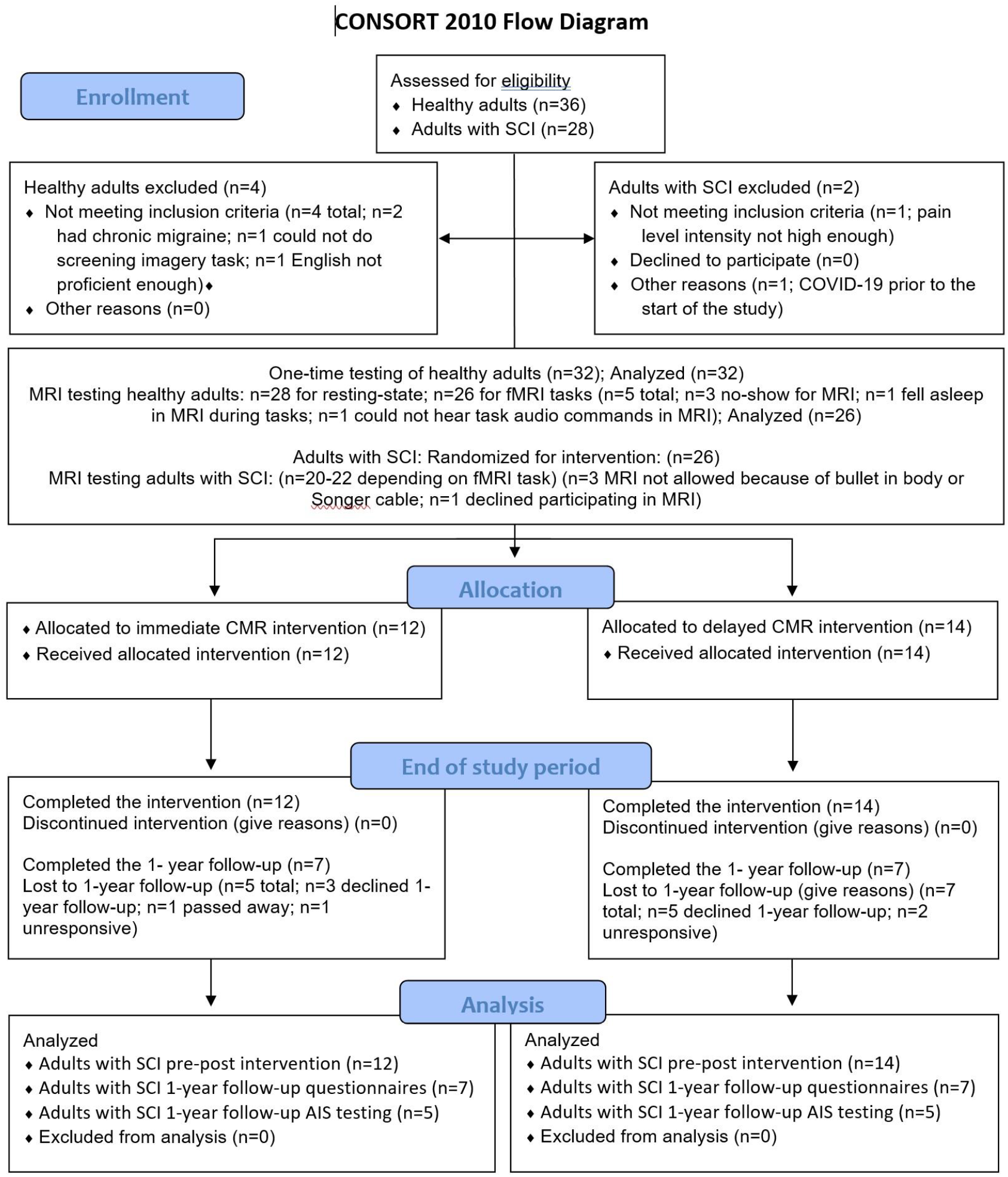
CONSORT flow diagram.

All 26 adults with SCI completed the intervention (18 sessions) and pre-post assessments. There were no study-related adverse events. They were 52±15 years of age, 1-56 years post-SCI, 14 participants with complete paraplegia, eight with incomplete paraplegia, two with incomplete tetraplegia, and two with Brown-Sequard. There were seven women (27%), two participants of Hispanic background (8%), one of Asian descent (4%), one of African American/Black descent (4%), and three participants living in a rural location (12%). The gender identity matched the biological sex allocation in all participants. About half of the participants used a manual wheelchair (46%) or a combination of manual and power wheelchair (12%). About 19% used a power wheelchair 100% of the time and about 23% of the participants walked with or without assistance.

Their highest baseline neuropathic pain of 7.81±1.33 on the NPRS was reduced to 2.88±2.92 after 6 weeks of CMR and nine participants were pain-free after the intervention (34.62%). Of the 14 participants that completed the 1-year follow-up, four were still completely pain-free, four other participants were still at a 0 for average pain, four remained at the reduced pain level they had at 6-week follow-up (with average pain of 2 out of 10 on the NPRS), and only two participants scored respectively 5 and 6 on average pain, related to infections or surgeries at 1-year follow-up, causing a temporary increase in highest and average neuropathic pain levels.

***Table 2*** shows the means and standard deviations of the most relevant clinical assessments across all time points as well as the within- and between-group statistical analysis. We found significant differences between group A (early CMR) and group B (observation period) for highest, average, lowest neuropathic pain, average nociceptive pain, and INSCI AIS exam of sensation (touch, pin prick) and lower limb motor function. Group A improved in sensorimotor function and had significant neuropathic pain reduction after 6 weeks of CMR, whereas group B (***Figure 2, line graph on the left)*** did not show any reductions in neuropathic pain and did not have any changes in sensation or motor function (INSCI ASIA exam) during the observation period (***Figure 3, bar graph on the left)***. Interference of neuropathic pain on activity, mood, and sleep, as well as average neuropathic and nociceptive pain, did not change during this period (***Figure 4, top figure***).

**Table 2.** Brain imaging outcomes for resting-state and task-based fMRI in healthy adults and adults with SCI.

|  | Z score | p-value<br>(FEW-corrected) | Cluster<br>extent<br>(voxels) | x (mm) | y (mm) | z (mm) |
| --- | --- | --- | --- | --- | --- | --- |
| Baseline Resting-state functional connectivity (right insula seed)<br>Healthy controls > Adults with SCI |  |  |  |  |  |  |
| Cluster 1 | 4.3 | <0.0001 | 14934 | -40 | 6 | 28 |
| Cluster 2 | 4.43 | 0.0007 | 2436 | 60 | -50 | -24 |
| Baseline Resting-state functional connectivity (right parietal operculum seed)<br>Healthy controls > Adults with SCI |  |  |  |  |  |  |
| Cluster 1 | 4.42 | <0.0001 | 14792 | -32 | -40 | 54 |
| Baseline Sensation Right Toe<br>Healthy controls > Adults with SCI |  |  |  |  |  |  |
| Cluster 1 | 5.84 | <0.0001 | 1922 | -2 | -24 | 70 |
| Cluster 2 | 4.66 | <0.0001 | 1039 | -62 | -20 | 16 |
| Cluster 3 | 4.64 | <0.0001 | 595 | -60 | 14 | 38 |
| Cluster 4 | 4.08 | 0.0005 | 374 | -58 | -22 | 44 |
| Cluster 5 | 3.95 | 0.0027 | 326 | 52 | -30 | 20 |
| Adults with SCI: Resting-state functional connectivity - parietal operculum seed, Post>Pre CMR |  |  |  |  |  |  |
| Cluster 1 | 3.83 | 0.0125 | 682 | -46 | -34 | 26 |
| Adults with SCI: Sensation-right toe, Post > Pre CMR |  |  |  |  |  |  |
| Cluster 1 | 3.48 | <0.0001 | 1428 | -34 | -36 | 42 |
| Cluster 2 | 3.4 | <0.0001 | 1341 | 22 | -66 | 52 |
| Cluster 3 | 3.53 | <0.0001 | 775 | -34 | 22 | -6 |
| Cluster 4 | 3.41 | <0.0001 | 511 | -22 | -14 | 60 |
| Cluster 5 | 3.73 | 0.0002 | 466 | -52 | 16 | 46 |

**Fig. 2.**
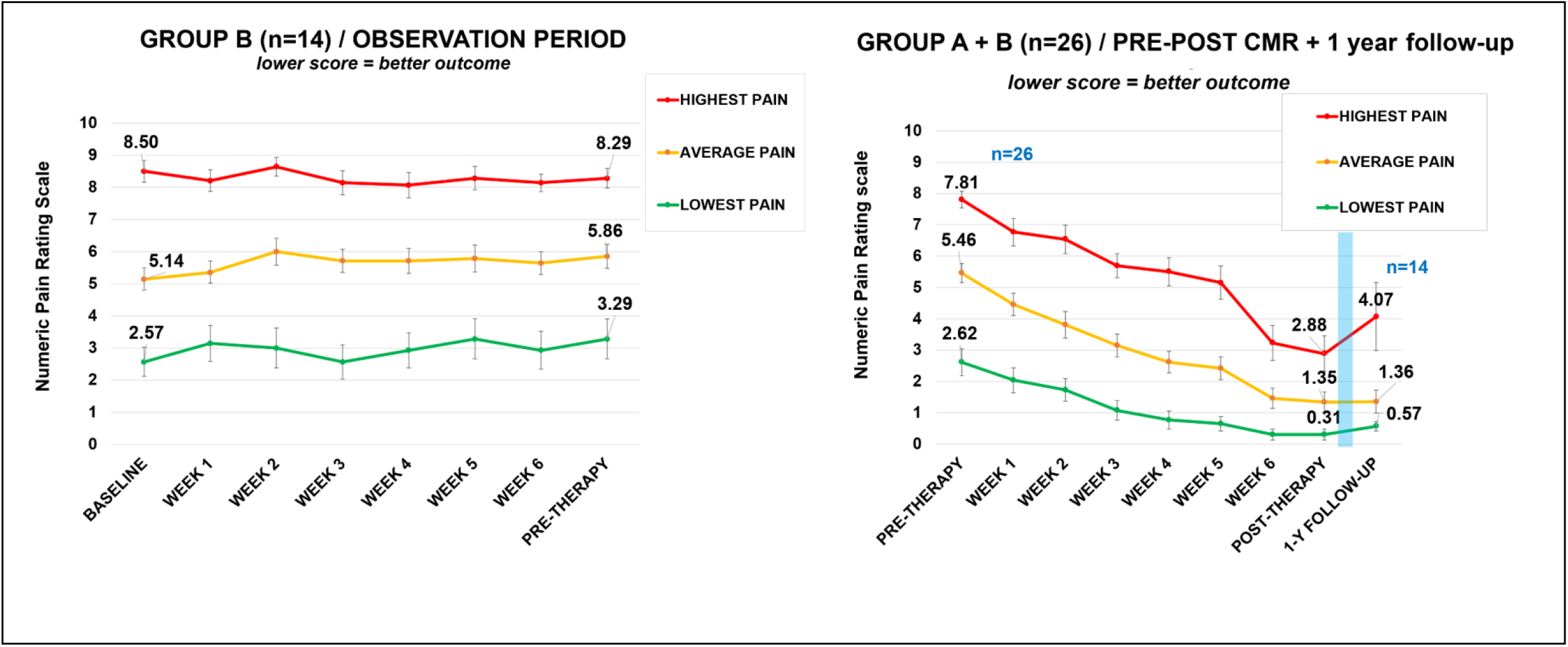
Highest, average, and lowest neuropathic pain rating scores. The line graphs represent the weekly neuropathic pain intensity ratings during the 6-week observation period (Group B, n=14, in the first 6 weeks, graph on the left) and during the pre-post CMR intervention (group A + B receiving CMR, n=26, graph on the right) with the one-year follow-up testing included (n=14).

**Fig. 3.**
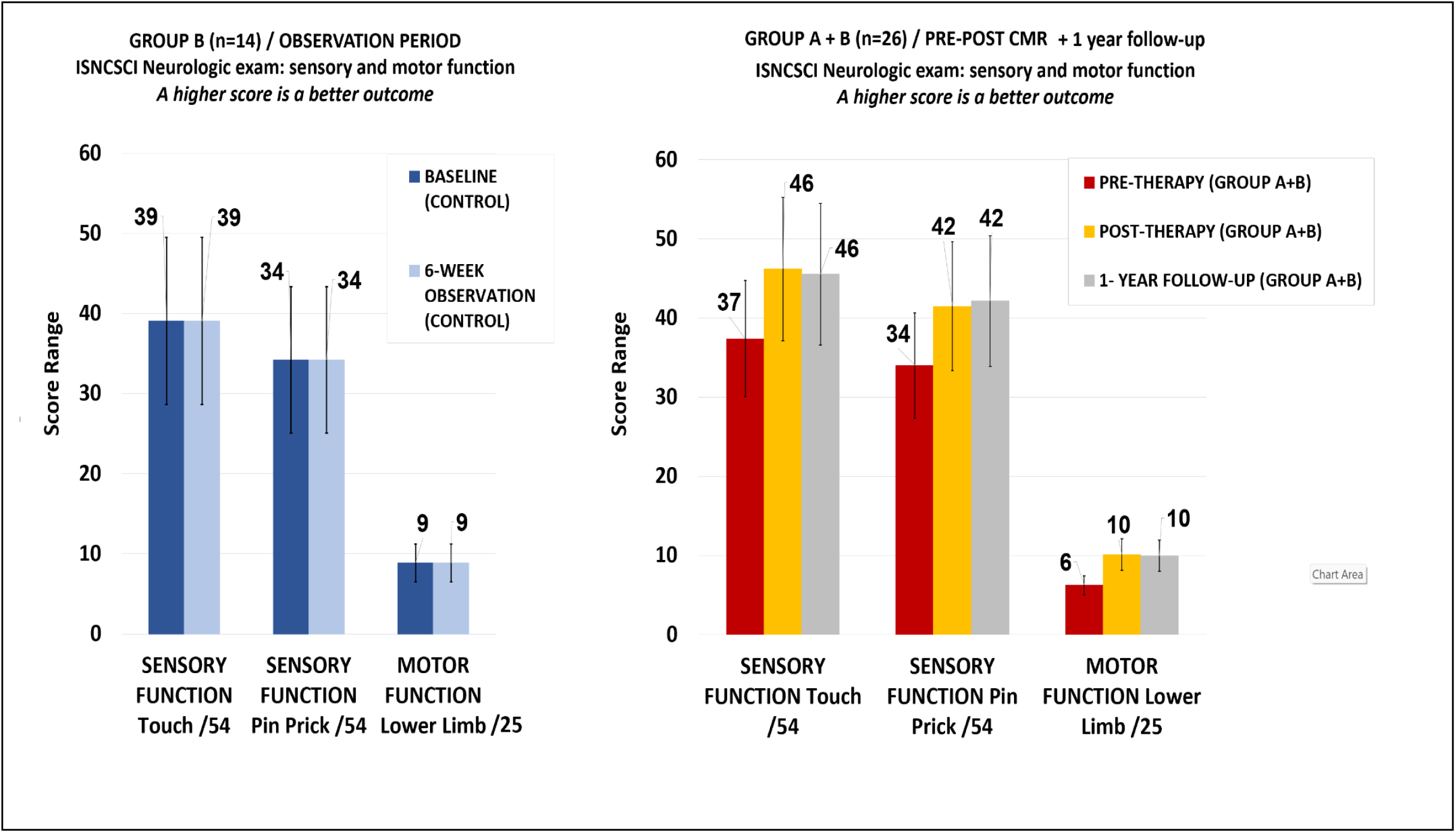
INSCI AIS exam. The bar graphs represent the touch and pin prick sensation as well as lower limb motor function during the 6-week observation period (Group B, n=14, in the first 6 weeks, bar graph on the left) and during the pre-post CMR intervention (group A + B receiving CMR, n=26, bar graph on the right) with the one-year follow-up testing included (n=14).

**Fig. 4.**
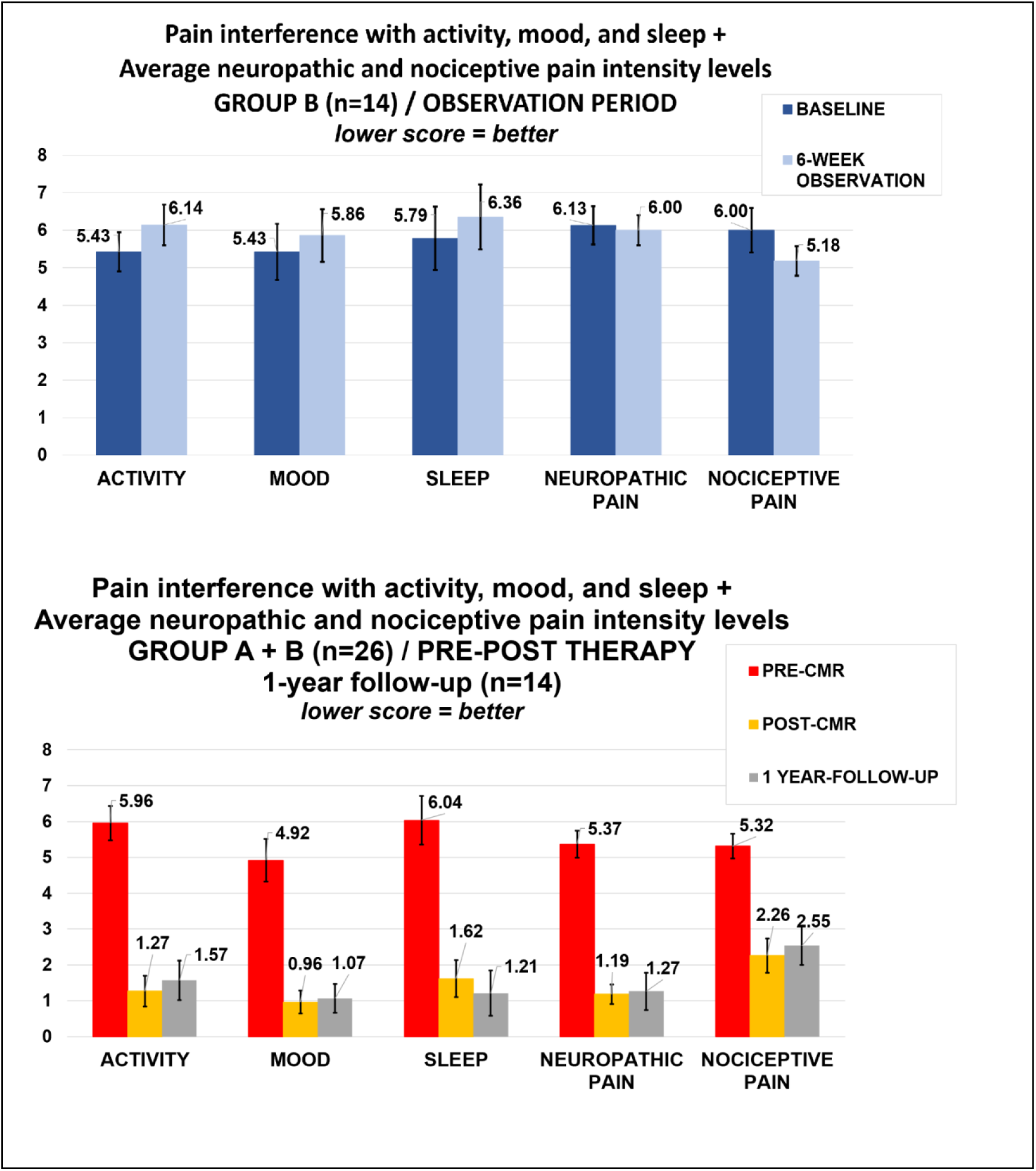
NINDS-CDE International SCI Pain Basic Data Set Version 2.0. Interference of neuropathic pain with activity, mood, and sleep, average neuropathic pain levels, average nociceptive (musculoskeletal) pain intensity levels at baseline and after the 6-week observation period (Group B, n=14, in the first 6 weeks, bar graph on the top) and pre-post CMR intervention, and at the 1-year follow-up (group A + B receiving CMR, n=26, bar graph on the bottom).

The within-group analysis of pre-post CMR intervention (group A and B combined, ***Figure 2, line graph on the right***) showed significant reductions in highest neuropathic pain (change score of, 4.92±2.92, large effect size Cohen’s *d*=1.68), average pain (change score 4.12±2.23, *d*=1.85), lowest pain (change score 2.31±2.07, d=1.00). A significant reduction of interference of neuropathic pain on activity, mood, and sleep was seen, as well as a significant reduction in average neuropathic and nociceptive pain. These improvements were also maintained at 1-year follow-up (***Figure 4, bottom figure***).

Significant improvements on the INSCI AIS exam were also seen after 6 weeks of CMR (group A and B combined, n=26, ***Figure 3, bar graph on the right***), namely improvements of touch sensation by 8.81±5.37 points (*d*=1.64), for pin prick sensation by 7.50±4.89 points (*d*=1.53), and for lower limb muscle strength by 3.87±2.81 points (*d*=1.38). The improvements were maintained at 1-year follow-up.

In addition, participants reported markedly improved function: twelve out of 20 adults previously unable to stand, could stand up for the first time with minimal assistance. Seventeen out of 18 adults with balance problems while sitting at baseline could lean forward and sideways confidently after the intervention. All participants reported they now felt the position of their legs in space, and the majority of them could feel the body weight distribution on the soles of their feet and the contact of the soles of the feet with the floor, facilitating weight shifts during transfers.

At baseline, all fMRI tasks, except the pain imagery, elicited activation in our areas of interest: pre-or postcentral gyrus (important for sensorimotor function), parietal operculum and insula (important for pain processing, and body awareness), and areas in the posterior parietal cortex (superior parietal lobe, angular gyrus, supramarginal gyrus) related to the creation of visuospatial body maps at each moment in time to guide movements.^46,64,68,70,71,100–102^ The pain imagery task mainly elicited activation in the frontal pole and the frontal orbital cortex. The frontal pole plays an important role for cognitive abilities such as action selection, working memory, and perception.^103,104^ The orbitofrontal cortex receives input from the five senses and from bodily signals, including the visceral sensory system.^105,106^ This brain area also has direct reciprocal connections with other brain structures, such as the insula and parietal operculum.^107^. A reason why the pain imagery elicited different areas than other tasks, might be related to the way the task was set up. Participants focused on their own highest pain location, and this was in a different body location for each person. Therefore, brain activations common to the whole group would be harder to find, whereas activation of brain areas related to attention to bodily signals would be expected and was found.

The most important brain imaging results, related to comparisons between healthy adults and adults with SCI, as well as changes in SCI after the CMR intervention are detailed in ***Table 3***. We demonstrated that the parietal operculum as well as insula networks have weaker connections in 20 adults with SCI-related neuropathic pain compared to 28 healthy adults. (***Figure 5***). Specific weaker connections were found between the right insula and the left frontal operculum, left insula, left parietal operculum (OP1/OP4), left angular gyrus, left supramarginal gyrus, and left superior parietal lobe. The frontal operculum and insula are related to body ownership.^108^ Interestingly, the intensity of chronic back pain has also been encoded in both these brain areas.^109^ Similarly, connectivity strength was weaker in adults with SCI compared to healthy adults between the right parietal operculum (ROI) and the left parietal operculum, left angular gyrus, and left supramarginal gyrus. Both resting-state connectivity results are in line with our model for altered sensorimotor integration and mental body representation deficits (i.e., body awareness/visuospatial body maps) in adults with SCI.

**Table 3.**
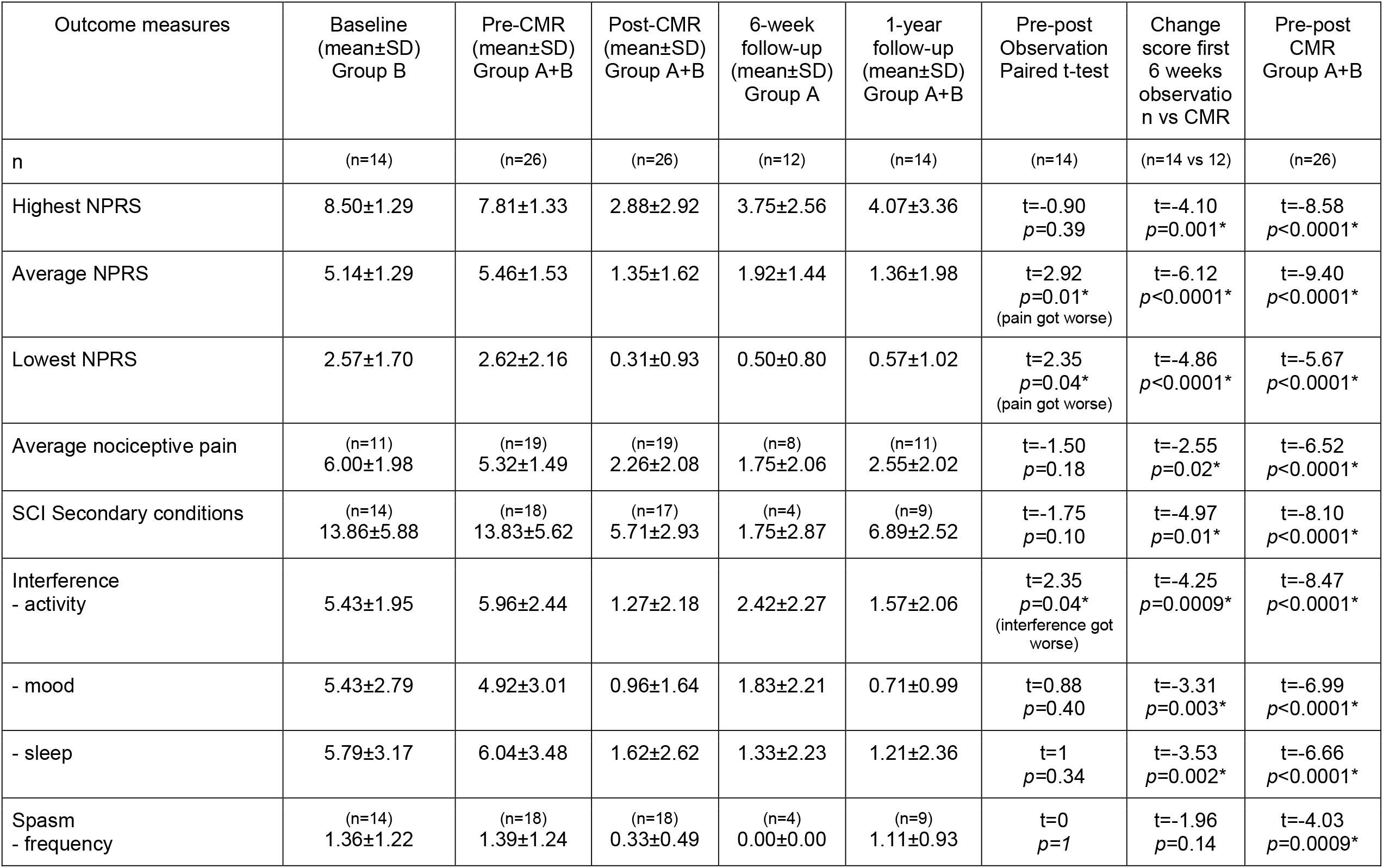
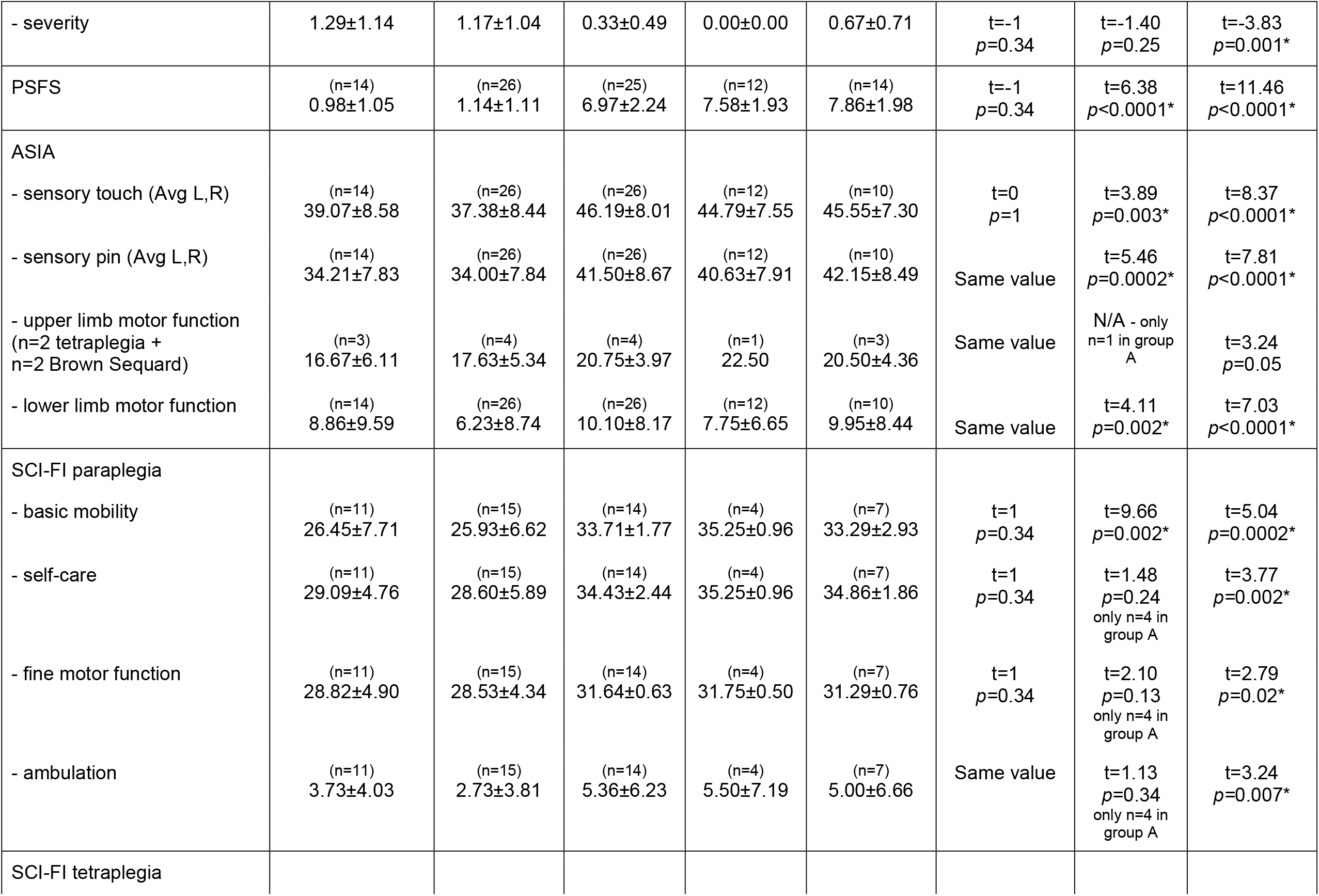
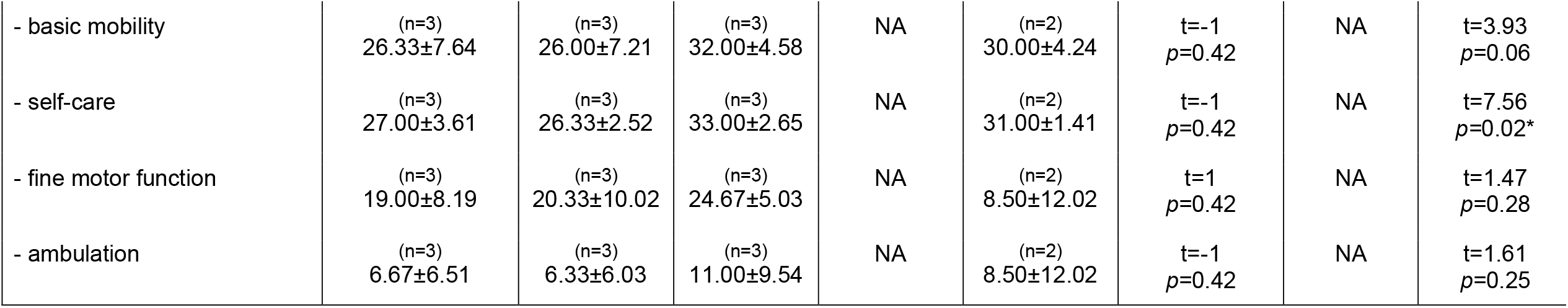
Primary and secondary outcome measures at five time points.

**Fig. 5.**
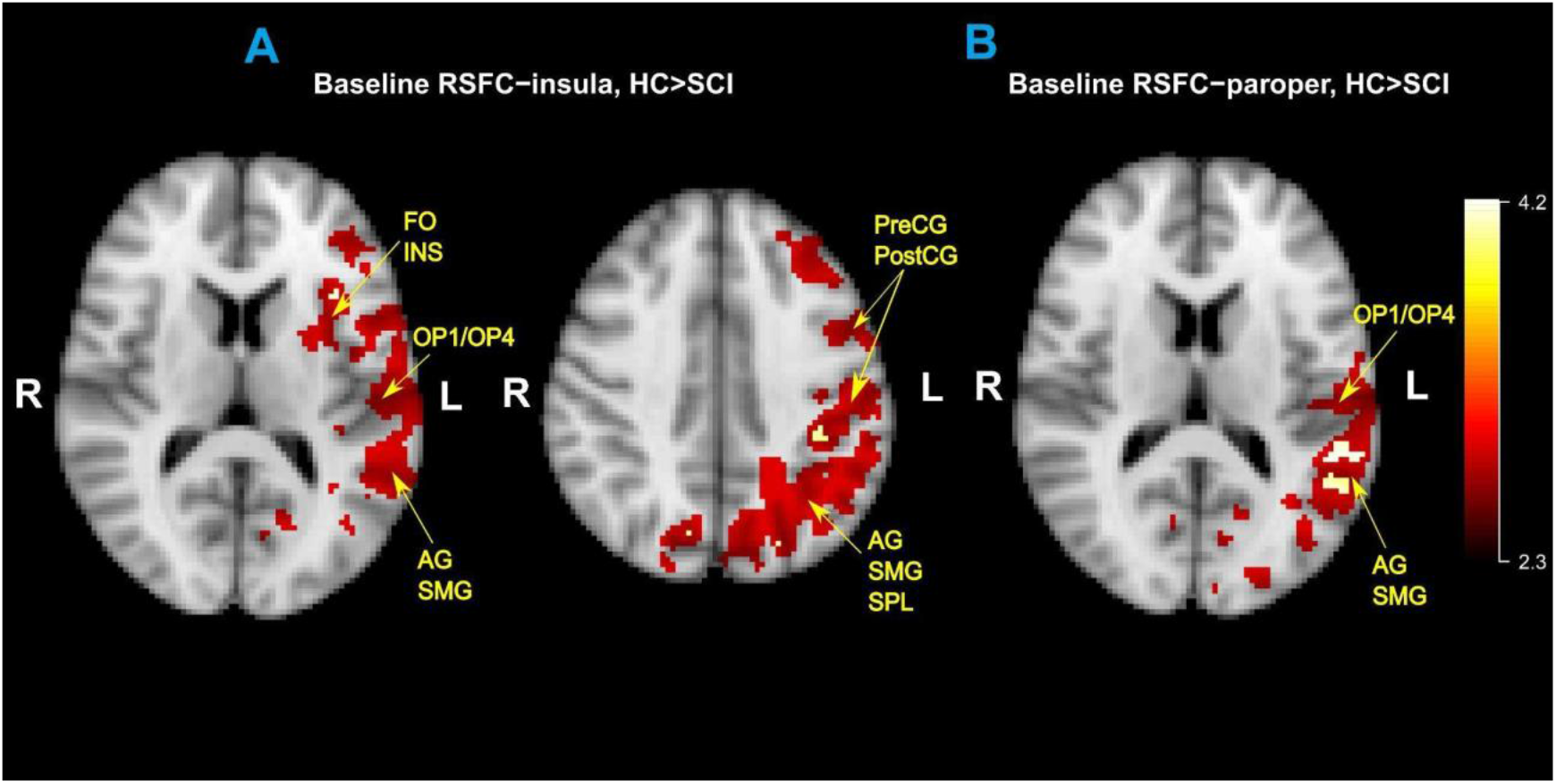
Resting-state functional connectivity baseline differences between 28 healthy adults vs 20 adults with SCI-related neuropathic pain. The two brain images on the left represent the insula network and the image on the right represents the parietal operculum network. The Region-of-Interest (ROI) seeds (right parietal operculum and right insula) have weaker connectivity with brain areas in the left hemisphere in adults with SCI compared to healthy adults. Legend: AG: angular gyrus; INS: insula; FO: frontal operculum; OP1/OP4: Parts 1 and 4 of the parietal operculum; preCG: precentral gyrus; postCG: postcentral gyrus; SMG: supramarginal gyrus; SPL: superior parietal lobe

Furthermore, at baseline, right toe sensory stimulation elicited less brain activation in 22 adults with SCI compared to 26 healthy adults (***Figure 6***). The areas that were less activated were the left pre- and post-central gyrus (congruent with what would be expected with right toe stimulation), and the bilateral parietal operculum, left supramarginal gyrus, and left superior parietal lobe. This result was expected given that the majority of our participants with SCI (n=14 out of 22 participants or 64%) could not feel when their toes were gently brushed with a small towel. In those that had some sensation, toe sensation was altered in three participants (13%), meaning that they either felt some tingling in the feet or located the sensation in the wrong place of the foot (e.g., heel), three other participants (13%) felt the toe sensation in only one foot, and only two participants (9%) could feel the toe sensation at baseline.

**Fig. 6.**
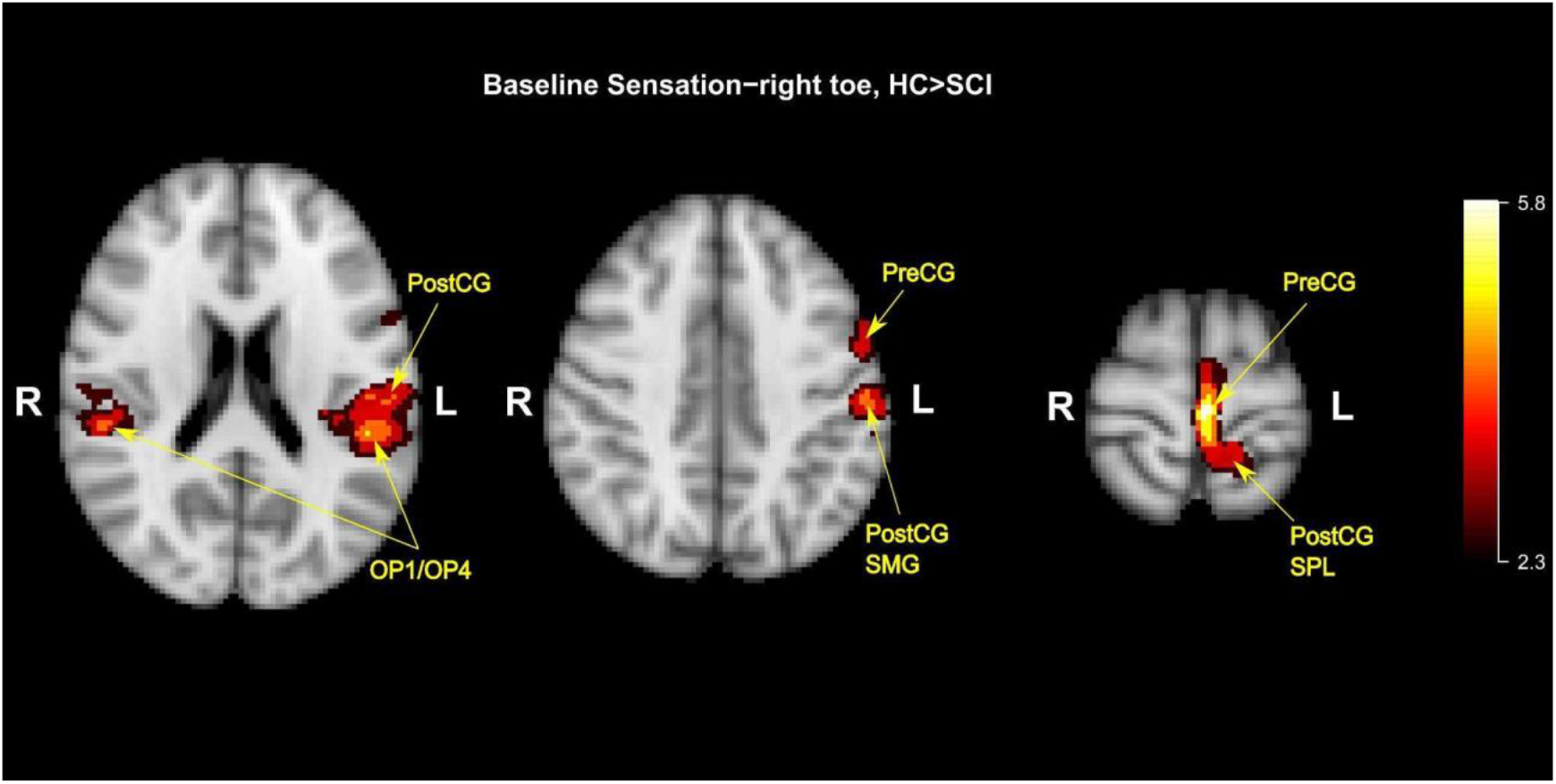
Baseline differences between 26 healthy adults vs 22 adults with SCI-related neuropathic pain for fMRI task of right toe sensory stimulation. Less brain activation in adults with SCI compared to healthy adults is seen in sensorimotor areas (left precentral gyrus (preCG) and left postcentral gyrus (postCG)), and parietal areas important for body awareness (bilateral parietal operculum parts OP1/OP4) and the creation and maintenance of visuospatial body maps (left supramarginal gyrus (SMG), and left superior parietal lobe (SPL).

Our two most interesting brain imaging results were the following: First, a stronger parietal operculum network connectivity was seen between the right parietal operculum and the left parietal operculum, left angular gyrus, and left supramarginal gyrus after the 6-week CMR intervention in adults with SCI (***Figure 7A***). Second, after CMR, increased brain activation in relevant sensorimotor areas and parietal areas related to pain, body awareness, and the visuospatial body map was seen during the fMRI task of right toe sensory stimulation. The brain areas were the bilateral postcentral gyrus, bilateral angular gyrus, bilateral supramarginal gyrus, bilateral superior parietal lobe, left insula, and left frontal operculum (***Figure 7B***). In comparison with the results above, after CMR, seven participants could correctly identify sensation in both toes during the fMRI task (32%) as opposed to only 2 participants (9%) at baseline. Four participants had altered sensation (18%); four other participants could correctly identify the sensation in one foot, but not the other (18%); and seven participants did not have sensation in the feet (32%) after CMR, which is 50% fewer participants with absent foot sensation compared to baseline. These brain imaging results were accompanied by AIS reports of improved touch sensation in the lower limbs (improvement of 8.81±5.37 points, *d*=1.6). The majority of participants reported they could now feel their feet in daily life, how the feet are positioned on the foot plates of the wheelchair (and not falling off anymore), and how the weight of the body is shifting to the soles of their feet during transfers, which increased their confidence to put weight on their legs, thereby improving the flow and speed of those transfers.

**Fig. 7.**
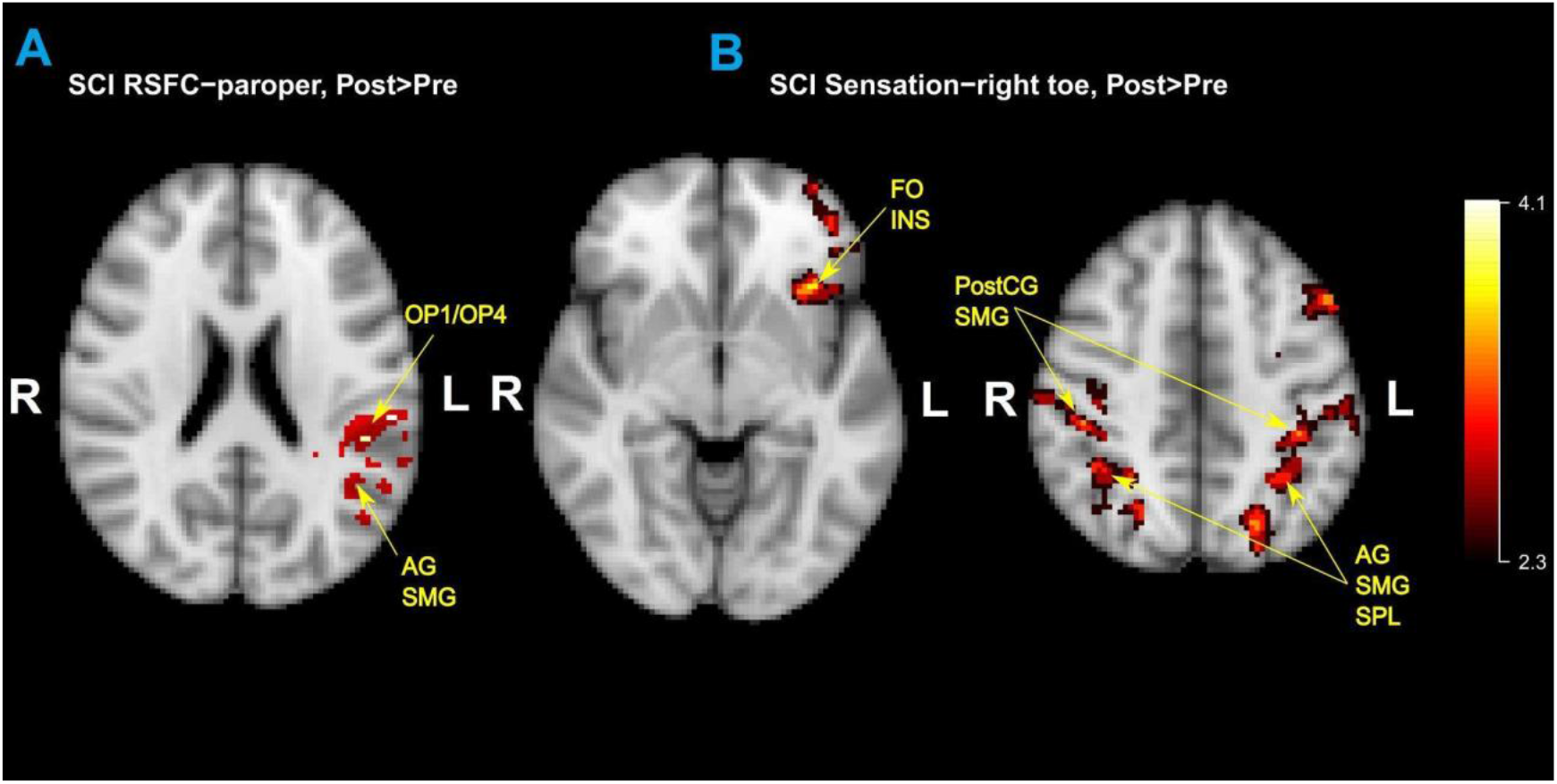
Pre-post CMR intervention differences: (**A**) A stronger parietal operculum network connectivity is seen in adults with SCI (n=20) after 6 weeks of CMR with stronger connections between the right parietal operculum (ROI) and the left parietal operculum (parts OP1/OP4), left angular gyrus (AG), and left supramarginal gyrus (SMG). (**B**) Increased task-related activation is seen for right toe sensory stimulation in adults with SCI-related neuropathic pain (n=21) after the CMR intervention, in line with increased sensation, evaluated with AIS testing. The activation was stronger in bilateral sensorimotor areas and areas important for body awareness and visuospatial body maps. Legend: AG: angular gyrus; INS: insula; FO: frontal operculum; postCG: postcentral gyrus; SMG: supramarginal gyrus; SPL: superior parietal lobe.

## 4. DISCUSSION

The aims of this phase I RCT were (1) to determine baseline differences in resting-state and task-based fMRI brain function in adults with SCI compared to healthy controls and (2) to identify changes in brain function and behavioral pain outcomes in adults with SCI after CMR. The goal of CMR is to restore awareness of the paralyzed limbs and trunk in space in areas of impaired sensorimotor function and/or neuropathic pain in order to relieve pain and improve sensorimotor function. Given the importance of sensory feedback for movement, sensory improvements create the opportunity for motor recovery to occur.

Our results confirmed our hypothesis that connectivity in parietal operculum and insula networks is weaker in adults with SCI compared to healthy adults, thereby expanding the knowledge of earlier work on altered structural and functional brain activity and connectivity in adults with SCI.^6,30,54,55^

Our most important results were the significant neuropathic pain relief and functional recovery in adults with SCI after 6 weeks of CMR, which we attributed to increased body awareness, and recovery of sensation and movement after CMR. These findings were coupled with evidence of brain function and connectivity restoration. The results indicate the preliminary efficacy of CMR for restoring function in adults with chronic SCI. Moreover, similar to our earlier study in stroke,^64^ we found that the parietal operculum network connectivity was stronger after 6 weeks of CMR in adults with SCI-related neuropathic pain, confirming hereby that focusing on restoring body awareness has a modulating effect on this network that is important for both pain processing and body awareness.^44^ The results further confirm the hypothesis that normalizing pain perception through restoring OP1/OP4 connectivity in adults with SCI can relieve neuropathic pain.

Limitations in the study were the limited racial and ethnic diversity in our sample. We also did not have Veterans in our sample, and only 12% of our participants were living in a rural location. A remote version of CMR could make this therapy accessible for patients who live in rural, underserved areas. CMR (in-person or remotely delivered) could also be beneficial for Veterans, given that 26% of adults with SCI receive care in a Veterans Health Administration (VA) setting.

So far, therapies have had limited success in neuropathic pain relief and functional recovery in adults with chronic SCI. CMR has shown preliminary efficacy to alleviate neuropathic pain relief and to improve function. The present results will inform and justify larger efficacy randomized clinical trials. We will use our existing community connections at the University of Minnesota to disseminate information and spread awareness of CMR and its results, design future studies, seek additional funding, and educate consumers, caregivers, and providers locally and nationally. CMR is easily implementable in current physical therapy practice. As part of the dissemination and clinical implementation, the first author (AVDW) and her team are conducting introduction and certification CMR classes at the University of Minnesota, in collaboration with the center in Italy (Centro Studi di Riabilitazione Neurocognitiva - Villa Miari (Study Center for Cognitive Multisensory Rehabilitation), to physical and occupational therapists, and doctors in the United States in order to make CMR accessible as a new rehabilitation approach in the United States.

## Data Availability

All data produced in the present study are available upon reasonable request to the authors.

## Declaration of interests

We declare having no competing interests.

## Acknowledgments

The AIRP2-IND-30: Academic Investment Research Program (AIRP) University of Minnesota School of Medicine funded this study (IRB no. STUDY00008476;ClinicalTrials.gov Identifier: NCT04706208). Additional support is provided by the National Center for Advancing Translational Sciences of the National Institutes of Health Award Number UL1TR002494, the Biotechnology Research Center: P41EB015894, the National Institute of Neurological Disorders & Stroke Institutional Center Core Grants to Support Neuroscience Research: P30 NS076408; and the High-Performance Connectome Upgrade for Human 3T MR Scanner: 1S10OD017974. Image processing resources were provided by the Minnesota Supercomputing Institute (MSI) at the University of Minnesota. The content is solely the responsibility of the authors and does not necessarily represent the official views of the National Institutes of Health.

We thank Dr. Marina Zernitz, Director of the Centro Studi di Riabilitazione Neurocognitiva - Villa Miari (Study Center for Cognitive Multisensory Rehabilitation), for her consultancy with the content of the cognitive multisensory rehabilitation training protocol. We would like to extend our profound thanks to Marc Noël for the critical review of the manuscript.

## Notes

### Competing Interest Statement

The authors have declared no competing interest.

### Clinical Trial

NCT04706208

### Clinical Protocols

https://doi.org/10.46292/sci22-00006

### Author Declarations

The University of Minnesota (UMN) Institutional Review Board approved the study (IRB #STUDY00008476).

## References

1 Taylor SM, Cheung EO, Sun R, Grote V, Marchlewski A, Addington EL. Applications of complementary therapies during rehabilitation for individuals with traumatic Spinal Cord Injury: Findings from the SCIRehab Project. J Spinal Cord Med 2019; 42: 571–8.

2 National Spinal Cord Injury Statistical Center. Spinal Cord Injury Facts and Figures at a Glance: 2022 SCI Data Sheet. https://msktc.org/sites/default/files/SCI-Facts-Figs-2022-Eng-508.pdf (accessed June 7, 2022).

3 Birmingham, AL: University of Alabama at Birmingham. Traumatic Spinal Cord Injury Facts and Figures at a Glance (2022 SCI Data Sheet). National Spinal Cord Injury Statistical Center (NSCISC). https://msktc.org/sites/default/files/SCI-Facts-Figs-2022-Eng-508.pdf (accessed June 6, 2022).

4 Leemhuis E, De Gennaro L, Pazzaglia AM. Disconnected Body Representation: Neuroplasticity Following Spinal Cord Injury. J Clin Med Res 2019; 8. DOI:10.3390/jcm8122144.

5 Lenggenhager B, Pazzaglia M, Scivoletto G, Molinari M, Aglioti SM. The sense of the body in individuals with spinal cord injury. PLoS One 2012; 7: e50757.

6 Leemhuis E, Giuffrida V, De Martino ML, et al. Rethinking the Body in the Brain after Spinal Cord Injury. J Clin Med Res 2022; 11. DOI:10.3390/jcm11020388.

7 Scandola M, Aglioti SM, Pozeg P, Avesani R, Moro V. Motor imagery in spinal cord injured people is modulated by somatotopic coding, perspective taking, and post-lesional chronic pain. Journal of Neuropsychology. 2017; 11: 305–26.

8 Kaur J, Ghosh S, Sahani AK, Sinha JK. Mental imagery training for treatment of central neuropathic pain: a narrative review. Acta Neurol Belg 2019; 119: 175–86.

9 Vastano R, Costantini M, Widerstrom-Noga E. Maladaptive reorganization following SCI: The role of body representation and multisensory integration. Prog Neurobiol 2022; 208: 102179.

10 Vastano R, Widerstrom-Noga E. Event-related potentials during mental rotation of body-related stimuli in spinal cord injury population. Neuropsychologia 2023; 179: 108447.

11 Vázquez-Fariñas M, Rodríguez-Martin B. ‘Living with a fragmented body’: a qualitative study on perceptions about body changes after a spinal cord injury. Spinal Cord 2021; 59: 855–64.

12 Osinski T, Martinez V, Bensmail D, Hatem S, Bouhassira D. Interplay between body schema, visuospatial perception and pain in patients with spinal cord injury. Eur J Pain 2020; 24: 1400–10.

13 Moro V, Corbella M, Ionta S, Ferrari F, Scandola M. Cognitive Training Improves Disconnected Limbs’ Mental Representation and Peripersonal Space after Spinal Cord Injury. Int J Environ Res Public Health 2021; 18. DOI:10.3390/ijerph18189589.

14 Moro V, Scandola M, Aglioti SM. What the study of spinal cord injured patients can tell us about the significance of the body in cognition. Psychon Bull Rev 2022; 29: 2052–69.

15 Maggio MG, Naro A, De Luca R, et al. Body Representation in Patients with Severe Spinal Cord Injury: A Pilot Study on the Promising Role of Powered Exoskeleton for Gait Training. J Pers Med 2022; 12. DOI:10.3390/jpm12040619.

16 Mehling WE, Wrubel J, Daubenmier JJ, et al. Body Awareness: a phenomenological inquiry into the common ground of mind-body therapies. Philos Ethics Humanit Med 2011; 6: 6.

17 De Martino ML, De Bartolo M, Leemhuis E, Pazzaglia M. Rebuilding Body–Brain Interaction from the Vagal Network in Spinal Cord Injuries. Brain Sciences 2021; 11: 1084.

18 Cardini F, Longo MR. Congruency of body-related information induces somatosensory reorganization. Neuropsychologia 2016; 84: 213–21.

19 Leemhuis E, Giuffrida V, Giannini AM, Pazzaglia M. A Therapeutic Matrix: Virtual Reality as a Clinical Tool for Spinal Cord Injury-Induced Neuropathic Pain. Brain Sci 2021; 11. DOI:10.3390/brainsci11091201.

20 Hatch MN, Cushing TR, Carlson GD, Chang EY. Neuropathic pain and SCI: Identification and treatment strategies in the 21st century. J Neurol Sci 2018; 384: 75–83.

21 Kramer JLK, Minhas NK, Jutzeler CR, Erskine ELK, Liu LJW, Ramer MS. Neuropathic pain following traumatic spinal cord injury: Models, measurement, and mechanisms. Journal of Neuroscience Research. 2017; 95: 1295–306.

22 Siddall PJ. Management of neuropathic pain following spinal cord injury: now and in the future. Spinal Cord. 2009; 47: 352–9.

23 Boldt I, Eriks-Hoogland I, Brinkhof MWG, de Bie R, Joggi D, von Elm E. Non-pharmacological interventions for chronic pain in people with spinal cord injury. Cochrane Database Syst Rev 2014; : CD009177.

24 Nardone R, Höller Y, Leis S, et al. Invasive and non-invasive brain stimulation for treatment of neuropathic pain in patients with spinal cord injury: a review. J Spinal Cord Med 2014; 37: 19–31.

25 Goel V, Kumar V, Patwardhan AM, et al. Procedure-Related Outcomes Including Readmission Following Spinal Cord Stimulator Implant Procedures: A Retrospective Cohort Study. Anesth Analg 2021; published online Dec 15. DOI:10.1213/ANE.0000000000005816.

26 Wang W, Xie W, Zhang Q, et al. Reorganization of the brain in spinal cord injury: a meta-analysis of functional MRI studies. Neuroradiology 2019; 61: 1309–18.

27 Solstrand Dahlberg L, Becerra L, Borsook D, Linnman C. Brain changes after spinal cord injury, a quantitative meta-analysis and review. Neurosci Biobehav Rev 2018; 90: 272–93.

28 Athanasiou A, Klados MA, Pandria N, et al. A Systematic Review of Investigations into Functional Brain Connectivity Following Spinal Cord Injury. Front Hum Neurosci 2017; 11: 517.

29 Jutzeler CR, Curt A, Kramer JLK. Relationship between chronic pain and brain reorganization after deafferentation: A systematic review of functional MRI findings. Neuroimage Clin 2015; 9: 599–606.

30 Black SR, King JB, Mahan MA, Anderson J, Butson CR. Functional Hyperconnectivity and Task-Based Activity Changes Associated With Neuropathic Pain After Spinal Cord Injury: A Pilot Study. Front Neurol 2021; 12: 613630.

31 Osinski T, Acapo S, Bensmail D, Bouhassira D. Central Nervous System Reorganization and Pain After Spinal Cord Injury: Possible Targets for Physical Therapy—A Systematic Review of Neuroimaging …. Physical 2020. https://academic.oup.com/ptj/article-pdf/doi/10.1093/ptj/pzaa043/33428407/pzaa043.pdf.

32 Oni-Orisan A, Kaushal M, Li W, et al. Alterations in Cortical Sensorimotor Connectivity following Complete Cervical Spinal Cord Injury: A Prospective Resting-State fMRI Study. PLoS One 2016; 11: e0150351.

33 Nees TA, Finnerup NB, Blesch A, Weidner N. Neuropathic pain after spinal cord injury: the impact of sensorimotor activity. Pain 2017; 158: 371–6.

34 Kaushal M, Oni-Orisan A, Chen G, et al. Evaluation of Whole-Brain Resting-State Functional Connectivity in Spinal Cord Injury: A Large-Scale Network Analysis Using Network-Based Statistic. J Neurotrauma 2017; 34: 1278–82.

35 Hanakawa T. Neural mechanisms underlying deafferentation pain: a hypothesis from a neuroimaging perspective. J Orthop Sci 2012; 17: 331–5.

36 Jutzeler CR, Freund P, Huber E, Curt A, Kramer JLK. Neuropathic Pain and Functional Reorganization in the Primary Sensorimotor Cortex After Spinal Cord Injury. J Pain 2015; 16: 1256–67.

37 Gustin SM, Wrigley PJ, Henderson LA, Siddall PJ. Brain circuitry underlying pain in response to imagined movement in people with spinal cord injury. Pain 2010; 148: 438–45.

38 Wrigley PJ, Press SR, Gustin SM, et al. Neuropathic pain and primary somatosensory cortex reorganization following spinal cord injury. Pain 2009; 141: 52–9.

39 Vuckovic A, Hasan MA, Fraser M, Conway BA, Nasseroleslami B, Allan DB. Dynamic oscillatory signatures of central neuropathic pain in spinal cord injury. J Pain 2014; 15: 645–55.

40 Finnerup NB, Norrbrink C, Trok K, et al. Phenotypes and predictors of pain following traumatic spinal cord injury: a prospective study. J Pain 2014; 15: 40–8.

41 Hawasli AH, Rutlin J, Roland JL, et al. Spinal Cord Injury Disrupts Resting-State Networks in the Human Brain. J Neurotrauma 2018; 35: 864–73.

42 Huynh V, Rosner J, Curt A, Kollias S, Hubli M, Michels L. Disentangling the Effects of Spinal Cord Injury and Related Neuropathic Pain on Supraspinal Neuroplasticity: A Systematic Review on Neuroimaging. Front Neurol 2019; 10: 1413.

43 Zhang Y-H, Hu H-Y, Xiong Y-C, et al. Exercise for Neuropathic Pain: A Systematic Review and Expert Consensus. Front Med 2021; 8: 756940.

44 Peyron R, Fauchon C. The posterior insular-opercular cortex: An access to the brain networks of thermosensory and nociceptive processes? Neurosci Lett 2019; 702: 34–9.

45 Garcia-Larrea L, Peyron R. Pain matrices and neuropathic pain matrices: A review. Pain. 2013; 154: S29–43.

46 Bretas RV, Taoka M, Suzuki H, Iriki A. Secondary somatosensory cortex of primates: beyond body maps, toward conscious self-in-the-world maps. Exp Brain Res 2020; 238: 259–72.

47 Friebel U, Eickhoff SB, Lotze M. Coordinate-based meta-analysis of experimentally induced and chronic persistent neuropathic pain. Neuroimage 2011; 58: 1070–80.

48 Hassanpour MS, Simmons WK, Feinstein JS, et al. The Insular Cortex Dynamically Maps Changes in Cardiorespiratory Interoception. Neuropsychopharmacology 2018; 43: 426–34.

49 Van de Winckel A, De Patre D, Rigoni M, et al. Exploratory study of how Cognitive Multisensory Rehabilitation restores parietal operculum connectivity and improves upper limb movements in chronic stroke. Sci Rep 2020; 10: 20278.

50 Van de Winckel A, Sunaert S, Wenderoth N, et al. Passive somatosensory discrimination tasks in healthy volunteers: Differential networks involved in familiar versus unfamiliar shape and length discrimination. NeuroImage. 2005; 26: 441–53.

51 Van de Winckel A, Wenderoth N, De Weerdt W, et al. Frontoparietal involvement in passively guided shape and length discrimination: a comparison between subcortical stroke patients and healthy controls. Exp Brain Res 2012; 220: 179–89.

52 Van de Winckel A, Klingels K, Bruyninckx F, et al. How does brain activation differ in children with unilateral cerebral palsy compared to typically developing children, during active and passive movements, and tactile stimulation? An fMRI study. Res Dev Disabil 2013; 34: 183–97.

53 Van de Winckel A, Verheyden G, Wenderoth N, et al. Does somatosensory discrimination activate different brain areas in children with unilateral cerebral palsy compared to typically developing children? An fMRI study. Res Dev Disabil 2013; 34: 1710–20.

54 Huynh V, Lütolf R, Rosner J, et al. Supraspinal nociceptive networks in neuropathic pain after spinal cord injury. Hum Brain Mapp 2021; 42: 3733–49.

55 Li X, Wang L, Chen Q, et al. The Reorganization of Insular Subregions in Individuals with Below-Level Neuropathic Pain following Incomplete Spinal Cord Injury. Neural Plast 2020; 2020: 2796571.

56 Perfetti C, Pante F, Rizzello C, et al. Il dolore come problema riabilitativo. Padova, Italia: Piccin, 2015.

57 Van de Winckel A, Sunaert S, Wenderoth N, et al. Passive somatosensory discrimination tasks in healthy volunteers: differential networks involved in familiar versus unfamiliar shape and length discrimination. Neuroimage 2005; 26: 441–53.

58 Lee S, Bae S, Jeon D, Kim KY. The effects of cognitive exercise therapy on chronic stroke patients’ upper limb functions, activities of daily living and quality of life. J Phys Therapy Sci 2015; 27: 2787–91.

59 Chanubol R, Wongphaet P, Chavanich N, et al. A randomized controlled trial of Cognitive Sensory Motor Training Therapy on the recovery of arm function in acute stroke patients. Clin Rehabil 2012; 26: 1096–104.

60 Marzetti E, Rabini A, Piccinini G, et al. Neurocognitive therapeutic exercise improves pain and function in patients with shoulder impingement syndrome: a single-blind randomized controlled clinical trial. Eur J Phys Rehabil Med 2014; 50: 255–64.

61 Morreale M, Marchione P, Pili A, et al. Early versus delayed rehabilitation treatment in hemiplegic patients with ischemic stroke: proprioceptive or cognitive approach? Eur J Phys Rehabil Med 2016; 52: 81–9.

62 Sallés L, Martín-Casas P, Gironès X, Durà MJ, Lafuente JV, Perfetti C. A neurocognitive approach for recovering upper extremity movement following subacute stroke: a randomized controlled pilot study. J Phys Therapy Sci 2017; 29: 665–72.

63 De Patre D, Van de Winckel A, Panté F, et al. Visual and Motor Recovery After ‘Cognitive Therapeutic Exercises’ in Cortical Blindness: A Case Study. J Neurol Phys Ther 2017; 41: 164–72.

64 Van de Winckel A, De Patre D, Rigoni M, et al. Exploratory study of how Cognitive Multisensory Rehabilitation restores parietal operculum connectivity and improves upper limb movements in chronic stroke. Scientific Reports. 2020; 10. DOI:10.1038/s41598-020-77272-y.

65 Sepulcre J, Sabuncu MR, Yeo TB, Liu H, Johnson KA. Stepwise connectivity of the modal cortex reveals the multimodal organization of the human brain. J Neurosci 2012; 32: 10649–61.

66 Sepulcre J. Integration of visual and motor functional streams in the human brain. Neurosci Lett 2014; 567: 68–73.

67 Sepulcre J. Functional streams and cortical integration in the human brain. Neuroscientist 2014; 20: 499–508.

68 Daprati E, Sirigu A, Nico D. Body and movement: consciousness in the parietal lobes. Neuropsychologia 2010; 48: 756–62.

69 Park H-D, Blanke O. Coupling Inner and Outer Body for Self-Consciousness. Trends Cogn Sci 2019; 23: 377–88.

70 Dijkerman HC, de Haan EH. Somatosensory processes subserving perception and action. Behav Brain Sci 2007; 30: 189–201; discussion 201–39.

71 Moro V, Pacella V, Scandola M, et al. A fronto-insular-parietal network for the sense of body ownership. Cereb Cortex 2023; 33: 512–22.

72 Whitlock JR. Posterior parietal cortex. Curr Biol 2017; 27: R691–5.

73 Berlucchi G, Vallar G. The history of the neurophysiology and neurology of the parietal lobe. In: Handbook of clinical neurology. Elsevier, 2018: 3–30.

74 de Vignemont F. Body schema and body image—Pros and cons. Neuropsychologia 2010; 48: 669–80.

75 Razmus M. Body representation in patients after vascular brain injuries. Cogn Process 2017; 18: 359–73.

76 Vignemont F de, de Vignemont F. Taxonomies of Body Representations. Oxford Scholarship Online. 2017. DOI:10.1093/oso/9780198735885.003.0009.

77 Pitron V, Alsmith A, de Vignemont F. How do the body schema and the body image interact? Conscious Cogn 2018; 65: 352–8.

78 Van de Winckel A, Carpentier S, Deng W, et al. Identifying body awareness-related brain network changes after cognitive multisensory rehabilitation for reduced neuropathic pain in adults with spinal cord injury: Protocol of the pilot clinical trial. TSCIR 2022 (in press).

79 WMA declaration of Helsinki – ethical principles for medical research involving human subjects. https://www.wma.net/policies-post/wma-declaration-of-helsinki-ethical-principles-for-medical-research-involving-human-subjects/ (accessed Nov 15, 2021).

80 Hanley MA, Jensen MP, Ehde DM, et al. Clinically significant change in pain intensity ratings in persons with spinal cord injury or amputation. Clin J Pain 2006; 22: 25–31.

81 Folstein MF, Folstein SE, McHugh PR. ‘Mini-mental state’: a practical method for grading the cognitive state of patients for the clinician. J Psychiatr Res 1975; 12: 189–98.

82 Malouin F, Richards CL, Jackson PL, Lafleur MF, Durand A, Doyon J. The Kinesthetic and Visual Imagery Questionnaire (KVIQ) for Assessing Motor Imagery in Persons with Physical Disabilities: A Reliability and Construct Validity Study. Journal of Neurologic Physical Therapy. 2007; 31: 20–9.

83 Perfetti C, Wopfner-Oberleit S. Der hemiplegische Patient: kognitiv therapeutische Übungen. Pflaum, 1997.

84 Perfetti C. L’exercice thérapeutique cognitif pour la rééducation du patient hémiplégique. Masson, 2001.

85 De Patre D, de Winckel V, Panté A, et al. A case report of visual and motor recovery after 8 months of ‘cognitive therapeutic exercises’ in cortical blindness. 2017.

86 Patriat R, Molloy EK, Meier TB, et al. The effect of resting condition on resting-state fMRI reliability and consistency: a comparison between resting with eyes open, closed, and fixated. Neuroimage 2013; 78: 463–73.

87 Wrigley PJ, Siddall PJ, Gustin SM. New evidence for preserved somatosensory pathways in complete spinal cord injury: A fMRI study. Hum Brain Mapp 2018; 39: 588–98.

88 Wahlgren C, Levi R, Amezcua S, Thorell O, Thordstein M. Prevalence of discomplete sensorimotor spinal cord injury as evidenced by neurophysiological methods: A cross-sectional study. J Rehabil Med 2021; 53: jrm00156.

89 Kirshblum S, Snider B, Rupp R, Read MS, International Standards Committee of ASIA and ISCoS. Updates of the International Standards for Neurologic Classification of Spinal Cord Injury: 2015 and 2019. Phys Med Rehabil Clin N Am 2020; 31: 319–30.

90 Widerström-Noga E, Biering-Sørensen F, Bryce TN, et al. The International Spinal Cord Injury Pain Basic Data Set (version 2.0). Spinal Cord 2014; 52: 282–6.

91 Mills PB, Vakil AP, Phillips C, Kei L, Kwon BK. Intra-rater and inter-rater reliability of the Penn Spasm Frequency Scale in People with chronic traumatic spinal cord injury. Spinal Cord 2018; 56: 569–74.

92 Kalpakjian CZ, Scelza WM, Forchheimer MB, Toussaint LL. Preliminary reliability and validity of a Spinal Cord Injury Secondary Conditions Scale. J Spinal Cord Med 2007; 30: 131–9.

93 Chu T. Spinal cord injury secondary conditions scale (SCI-SCS). SCIRE Professional. 2022; published online April 26. https://scireproject.com/outcome/spinal-cord-injury-secondary-conditions-scale-sciscs/ (accessed Feb 8, 2023).

94 Dragesund T, Strand LI, Grotle M. The Revised Body Awareness Rating Questionnaire: Development Into a Unidimensional Scale Using Rasch Analysis. Phys Ther 2018; 98: 122–32.

95 Spielberger CD. State-Trait Anxiety Inventory for Adults. DOI:10.1037/t06496-000.

96 Kroenke K, Spitzer RL, Williams JB. The PHQ-9: validity of a brief depression severity measure. J Gen Intern Med 2001; 16: 606–13.

97 Slavin MD, Ni P, Tulsky DS, et al. Spinal Cord Injury–Functional Index/Assistive Technology Short Forms. Arch Phys Med Rehabil 2016; 97: 1745–52.e7.

98 Westaway MD, Stratford PW, Binkley JM. The patient-specific functional scale: validation of its use in persons with neck dysfunction. J Orthop Sports Phys Ther 1998; 27: 331–8.

99 Keeney T, Slavin M, Kisala P, et al. Sensitivity of the SCI-FI/AT in Individuals With Traumatic Spinal Cord Injury. Arch Phys Med Rehabil 2018; 99: 1783–8.

100 Schellekens W, Bakker C, Ramsey NF, Petridou N. Moving in on human motor cortex. Characterizing the relationship between body parts with non-rigid population response fields. PLoS Comput Biol 2022; 18: e1009955.

101 van Stralen HE, Dijkerman HC, Biesbroek JM, et al. Body representation disorders predict left right orientation impairments after stroke: A voxel-based lesion symptom mapping study. Cortex 2018; 104: 140–53.

102 Dijkerman C, Lenggenhager B. The body and cognition: The relation between body representations and higher level cognitive and social processes. Cortex 2018; 104: 133–9.

103 Bludau S, Eickhoff SB, Mohlberg H, et al. Cytoarchitecture, probability maps and functions of the human frontal pole. Neuroimage 2014; 93 Pt 2: 260–75.

104 Kovach CK, Daw ND, Rudrauf D, Tranel D, O’Doherty JP, Adolphs R. Anterior prefrontal cortex contributes to action selection through tracking of recent reward trends. J Neurosci 2012; 32: 8434–42.

105 Kringelbach ML. The human orbitofrontal cortex: linking reward to hedonic experience. Nat Rev Neurosci 2005; 6: 691–702.

106 Critchley HD, Mathias CJ, Dolan RJ. Fear conditioning in humans: the influence of awareness and autonomic arousal on functional neuroanatomy. Neuron 2002; 33: 653–63.

107 Mesulam MM, Mufson EJ. Insula of the old world monkey. III: Efferent cortical output and comments on function. J Comp Neurol 1982; 212: 38–52.

108 Tsakiris M, Hesse MD, Boy C, Haggard P, Fink GR. Neural signatures of body ownership: a sensory network for bodily self-consciousness. Cereb Cortex 2007; 17: 2235–44.

109 Mayr A, Jahn P, Stankewitz A, et al. Patients with chronic pain exhibit individually unique cortical signatures of pain encoding. Hum Brain Mapp 2022; 43: 1676–93.

